# Understanding COVID-19 trajectories from a nationwide linked electronic health record cohort of 56 million people: phenotypes, severity, waves & vaccination

**DOI:** 10.1101/2021.11.08.21265312

**Authors:** Johan H Thygesen, Christopher Tomlinson, Sam Hollings, Mehrdad Mizani, Alex Handy, Ashley Akbari, Amitava Banerjee, Jennifer Cooper, Alvina Lai, Ken Li, Bilal Mateen, Naveed Sattar, Reecha Sofat, Ana Torralbo, Honghan Wu, Angela Wood, Jonathan A C Sterne, Christina Pagel, William Whiteley, Cathie Sudlow, Harry Hemingway, Spiros Denaxas, on behalf of the CVD-COVID-UK Consortium

## Abstract

**Background:** Updatable understanding of the onset and progression of individuals COVID-19 trajectories underpins pandemic mitigation efforts. In order to identify and characterize individual trajectories, we defined and validated ten COVID-19 phenotypes from linked electronic health records (EHR) on a nationwide scale using an extensible framework.

**Methods:** Cohort study of 56.6 million people in England alive on 23/01/2020, followed until 31/05/2021, using eight linked national datasets spanning COVID-19 testing, vaccination, primary & secondary care and death registrations data. We defined ten COVID-19 phenotypes reflecting clinically relevant stages of disease severity using a combination of international clinical terminologies (e.g. SNOMED-CT, ICD-10) and bespoke data fields; positive test, primary care diagnosis, hospitalisation, critical care (four phenotypes), and death (three phenotypes). Using these phenotypes, we constructed patient trajectories illustrating the transition frequency and duration between phenotypes. Analyses were stratified by pandemic waves and vaccination status.

**Findings:** We identified 3,469,528 infected individuals (6.1%) with 8,825,738 recorded COVID-19 phenotypes. Of these, 364,260 (11%) were hospitalised and 140,908 (4%) died. Of those hospitalised, 38,072 (10%) were admitted to intensive care (ICU), 54,026 (15%) received non-invasive ventilation and 21,404 (6%) invasive ventilation. Amongst hospitalised patients, first wave mortality (30%) was higher than the second (23%) in non-ICU settings, but remained unchanged for ICU patients. The highest mortality was for patients receiving critical care outside of ICU in wave 1 (51%). 13,083 (9%) COVID-19 related deaths occurred without diagnoses on the death certificate, but within 30 days of a positive test while 10,403 (7%) of cases were identified from mortality data alone with no prior phenotypes recorded. We observed longer patient trajectories in the second pandemic wave compared to the first.

**Interpretation:** Our analyses illustrate the wide spectrum of severity that COVID-19 displays and significant differences in incidence, survival and pathways across pandemic waves. We provide an adaptable framework to answer questions of clinical and policy relevance; new variant impact, booster dose efficacy and a way of maximising existing data to understand individuals progression through disease states.

**Research in Context:** *Evidence before the study:* We searched PubMed on October 14, 2021, for publications with the terms “COVID-19” or “SARS-CoV-2”, “severity”, and “electronic health records” or “EHR” without date or language restrictions. Multiple studies explore factors associated with severity of COVID-19 infection, and model predictions of outcome for hospitalised patients. However, most work to date focused on isolated facets of the healthcare system, such as primary or secondary care only, was conducted in subpopulations (e.g. hospitalised patients) of limited sample size, and often utilized dichotomised outcomes (e.g. mortality or hospitalisation) ignoring the full spectrum of disease. We identified no studies which comprehensively detailed severity of infections while describing disease severity across pandemic waves, vaccination status, and patient trajectories.

*Added value of this study:* To our knowledge, this is the first study providing a comprehensive view of COVID-19 across pandemic waves using national data and focusing on severity, vaccination, and patient trajectories. Drawing on linked electronic health record (EHR) data on a national scale (56.6 million people alive and registered with GP in England), we describe key demographic factors, frequency of comorbidities, impact of the two main waves in England, and effect of full vaccination on COVID-19 severities. Additionally, we identify and describe patient trajectory networks which illustrate the main transition pathways of COVID-19 patients in the healthcare system. Finally, we provide reproducible COVID-19 phenotyping algorithms reflecting clinically relevant stages of disease severity i.e. positive tests, primary care diagnoses, hospitalisation, critical care treatments (e.g. ventilatory support) and mortality.

*Implications of all the available evidence:* The COVID-19 phenotypes and trajectory analysis framework outlined produce a reproducible, extensible and repurposable means to generate national-scale data to support critical policy decision making. By modelling patient trajectories as a series of interactions with healthcare systems, and linking these to demographic and outcome data, we provide a means to identify and prioritise care pathways associated with adverse outcomes and highlight healthcare system ‘touch points’ which may act as tangible targets for intervention.

## Introduction

Understanding the population impact of COVID-19 requires consideration of how COVID-19 varies in severity, from asymptomatic to fatal, and time course, from acute infection to chronic sequelae termed ‘long COVID’. These diverse clinical manifestations are reflected in a patients’ digital trace across multiple, often unconnected, health system organisations including public health, general practice, hospitals, intensive care and civil death registration.

The trajectories of disease severity in COVID-19 are poorly understood for three reasons. Firstly, there is an important need for scale in order to identify outcomes in rare demographics, comorbidities and treatments. Secondly, there has been an unmet need to comprehensively link an individual’s data across currently siloed institutional datasets. In practice scale and linkage are intrinsically related concepts. In national health systems, as in England, datasets may encapsulate the population, yet be restricted to an isolated facet of the health system, e.g. primary care, meaning linkage is vital to capture the depth of patients’ interactions across aspects of healthcare. Conversely, in other health systems the unit of the dataset may be that of a single healthcare provider, encompassing rich patient data across both primary and secondary care, but for a limited population subset. In this later case linkage between providers becomes necessary to expand the breadth of individuals captured.

These issues are manifested in the existing literature where studies approaching a population-scale have for example been restricted to primary care^1^, and those reporting more detailed outcomes have been limited to population subsets^2^. Previous studies have attempted to use linkage to mitigate these issues, but have fallen short of reaching population-scale - e.g. Mathur et al. used five linked datasets to determine COVID-19 positive tests, hospital admissions, ICU and death^3^, however in using only a single dataset per health system this limits their study population (17.3 million individuals, ∼30% of English population) and event ascertainment.

Third, there is a need for open, reproducible electronic health record (EHR) COVID-19 phenotypes capturing significant treatments and outcomes such as intensive care admission and ventilatory support. Such phenotypes need to evolve to reflect changes in clinical practice and data recording, and incorporate uncertainty, e.g. clinical diagnoses in the absence of testing, or deaths occurring after infection, in the absence of COVID-19 as a documented cause.

Despite the proliferation of EHR-based COVID-19 research few studies have explicitly addressed phenotyping. Previous works have focussed on the development of binary phenotypes for COVID-19 infection and hospitalisation, by comparing a single terminology (ICD-10-CM) with test positivity^4^, or defining ‘severe COVID-19’ using laboratory tests, medications, diagnoses & procedures^5^. Neither of these approaches are directly transferable to English national EHR data and both fail to capture the full range of COVID-19 events including primary care and deaths.

To address these gaps, we leveraged nationwide data from England linking laboratory testing, primary care, hospitalisations (including critical care), and registered deaths to (a) define COVID-19 phenotypes reflecting clinically relevant stages of disease severity, (b) characterise demographics and comorbidities of those individuals experiencing these phenotypes, (c) compare disease severity, mortality and trajectories stratifying by pandemic wave, demographics and vaccination status. Through data linkage we establish an updatable framework able to reconstruct an individual’s COVID-19 trajectory across distinct severity states, providing vital insight which can be used to assess impact of new variants, booster dose efficacy, post-exposure prophylaxis and emerging drug treatments.

## Methods

### Study design and data sources

We conducted a cohort study using 8 linked National Health Service (NHS) datasets from the population of England available within the NHS Digital Trusted Research Environment (TRE) accessed through the CVD-COVID-UK / COVID-IMPACT Consortium. The following datasets were included: a) national laboratory COVID-19 testing data from the Public Health England (PHE) Second Generation Surveillance System (SGSS), b) primary care data from the General Practice Extraction Service Extract for Pandemic Planning and Research (GDPPR)^6^, c) hospital admission information from; Secondary Uses Service (SUS), Hospital Episode Statistics (HES) for Admitted Patient Care (HES-APC) and Adult Critical Care (HES-CC), d) COVID-19 hospitalisation information from COVID-19 Hospitalisations in England Surveillance System (CHESS), a dataset initiated by the NHS at the start of the pandemic to record information about patients hospitalised with COVID-19, e) COVID-19 Vaccination Status capturing vaccination details on a weekly basis, and f) mortality information from the Office for National Statistics (ONS) Civil Registration of Deaths^7^. Datasets were linked on an individual level by NHS Digital using a pseudonymised version of the NHS number, a unique 10 digit patient identifier used in the UK healthcare system.

A Reporting of studies Conducted using Observational Routinely-collected Data (RECORD) statement can be found in the supplement.

### Study population and pandemic waves

The study start date was 23rd January 2020, the date of the first recorded COVID-19 case in the UK^8^, and the end date was 31st May 2021. We included individuals that were: a) alive at the start of the study, b) registered with a General Practitioner (GP) in England (minimum one patient record in GDPPR), c) associated with a valid person pseudo-identifier enabling data linkage, d) had minimum 28 days of follow-up time, and e) residing in England, as defined using Lower-layer Super Output Areas (LSOA) (Supplementary Figure 1).

We defined pandemic waves using a data-driven approach, in the absence of a consensus definition. We defined the first wave as the period where more than 1,000 cases per day were reported by PHE (February 20th to May 29th 2020) and the second wave as the period with more than 10,000 cases per day (September 30th 2020 to February 12th 2021), accounting for the increase in testing capacity^9^. Individuals were assigned to waves based on the date of their first identified COVID-19 phenotype.

### Defining COVID-19 phenotypes

Our strategy for identifying COVID-19 from EHR spanning all healthcare settings relied on combining diagnosis codes, laboratory testing, disease outcomes and the provision of ventilatory support, both within and outside of the ICU. In order to improve the generalisability and reproducibility of our phenotypes, we created modular algorithms that can be adapted and applied in other datasets to make use of all available information for event ascertainment. We defined ten COVID-19 phenotypes reflecting clinically relevant stages of disease severity and encompassing five categories: a) positive tests, b) COVID-19 diagnosis recorded in primary care, c) hospital admissions, d) critical care (four phenotypes), and e) deaths (three phenotypes)(Figure 1 & Supplementary Table 1). Primary care diagnoses were ascertained using SNOMED-CT concepts while hospitalisations and relevant deaths were identified using ICD-10 terms.

**Figure 1:**
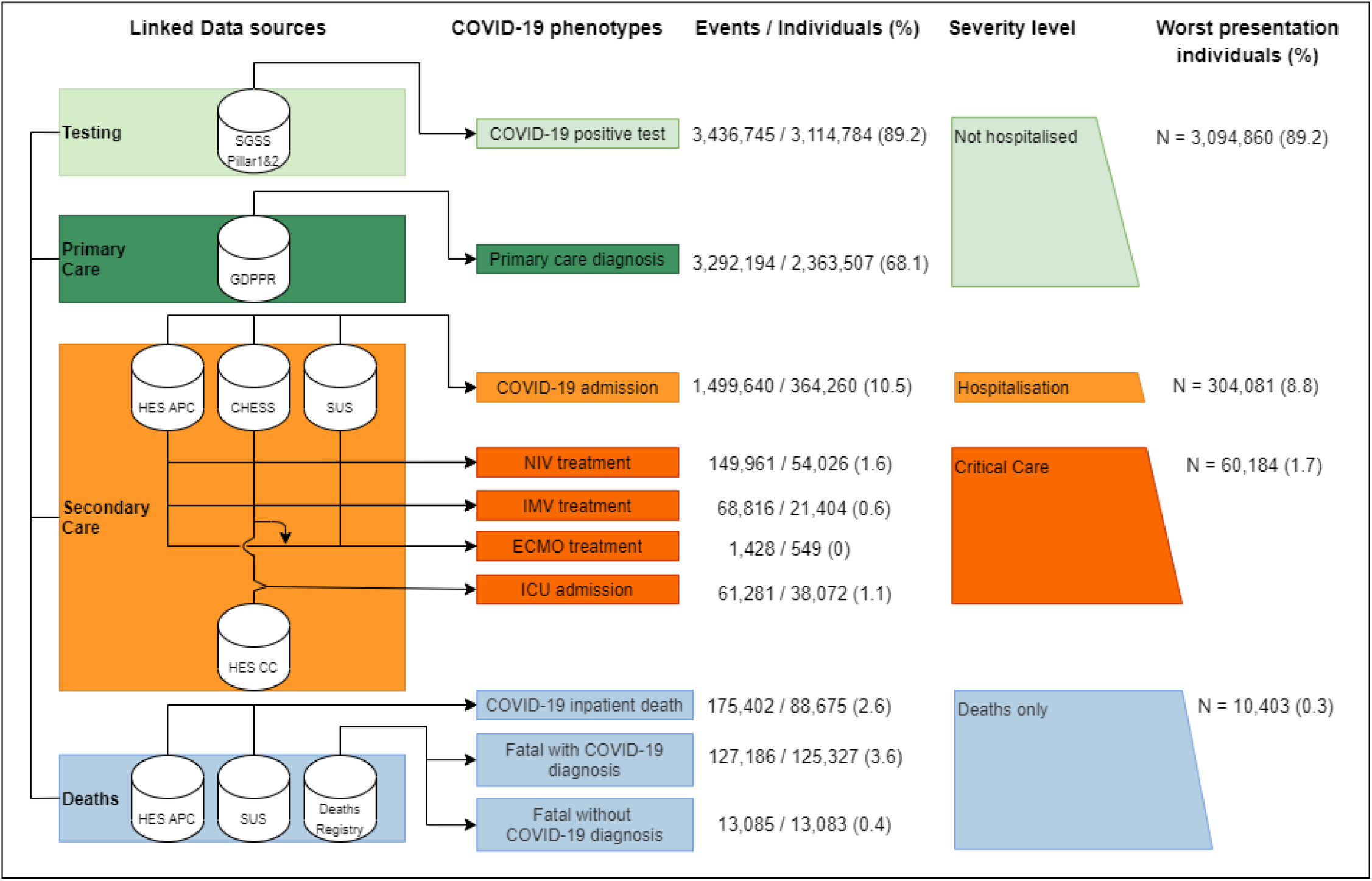
Framework describing the phenotyping of the ten COVID-19 phenotypes, and their severity categories, using seven linked data sources in 56.6 million people, to evaluate difference between wave and vaccination status. Sources used to identify each step are indicated with data buckets on the left and COVID-19 events in rectangles on the right. In all sources, ontology terms for both suspected and confirmed diagnosis were used. (%) indicate the percentage of individuals with a given COVID-19 event phenotype out of all individuals with any event phenotype. SGSS (Second Generation Surveillance System). Pillar 1 includes tests from NHS hospitals for those with a clinical need and healthcare workers. Pillar 2 is swab testing from the wider population. GDPPR (General Practice Extraction Service Extract for Pandemic Planning and Research) Primary care EHR diagnosis. HES (Hospital episode statistics), APC (Admitted Patient Care), CC (Critical Care), SUS (Secondary Uses Service) and CHESS (COVID-19 Hospitalisations in England Surveillance System). Ventilation support is defined either as NIV (Non-Invasive Ventilation), (IMV) Invasive Mechanical Ventilation or (ECMO) Extracorporeal Membrane Oxygenation (ECMO). HES CC does not give info on ECMO treatments.

We defined four critical care phenotypes based on ventilatory modalities, Non-Invasive Ventilation (NIV), Invasive Mechanical Ventilation (IMV), and Extracorporeal Membrane Oxygenation (ECMO), and Intensive Care Unit (ICU) admission. These phenotypes were defined using proprietary fields from HES-CC and CHESS alongside OPCS-4 procedure codes (analogous to Current Procedural Terminology (CPT) codes used in the US). Additional COVID-19 death phenotypes were created to ascertain not only deaths with COVID-19 as a named cause, but also deaths within 28 days of a positive test (inline with the PHE reported figures) and inpatient deaths.

We adopted the CALIBER rule-based phenotyping approach^10^ to create reproducible phenotypes which are shared publicly, alongside the study protocol and analytical code, on the HDR UK CALIBER Phenotype Library^11^ and GitHub^12^. Phenotype codelists are provided in Supplementary Table 2, and further details on phenotyping can be found in the method supplement.

### Demographics and comorbidities

Age, sex and ethnicity were derived from the most recent non-missing value across primary and secondary care (GDPPR, HES APC respectively), with preference to primary care in the event of a match on the same date. Ethnicity was categorised according to ONS census categories. Socioeconomic deprivation information was derived using the 2011 Lower-layer Super Output Areas (LSOA) from GDPPR to index the 2019 English indices of deprivation^13^ and IMD mapped to fifths (1 = most deprived, 5 = least).

We assessed 270 previously described comorbidities^14^, across 16 clinical specialities / organ systems, using validated CALIBER phenotypes and data records from 1st of January 1996 until 31st December 2019 from primary care, hospitalisation and procedure data^14^ (Supplementary Figure 3). A multimorbidity variable was created as the binary sum across all 270 conditions.

England has a policy of ‘Shielding’ uninfected patients who have specified underlying conditions that make them ‘clinically extremely vulnerable’ and at ‘very high’ risk of severe COVID-19 infections by advising them to remain at home^15^. Patients on the national Shielded Patient List, were identified by SNOMED-CT code ‘1300561000000107’ in their primary care records, used by GPs from 4th May 2020.

Vaccination status was determined from the COVID-19 vaccination dataset, including all vaccinations given after December 12th 2020 (when the first official dose was administered in England). Patients were denoted as vaccinated after 14 days had elapsed since their second dose. To examine effects, vaccinated patients were matched 1:1 with unvaccinated individuals on age (five year brackets), sex and ethnicity, drawing from individuals who had neither received a COVID-19 vaccine nor had previously ascertained COVID-19 infection. Analysis was performed from a single time point, February 1st 2021, with a minimum of 28 days follow-up.

### Statistical analyses

We used descriptive statistics to summarise patient populations and characteristics. Venn diagrams were used to illustrate the congruence of COVID-19 phenotype ascertainment between data sources.

COVID-19 event trajectory networks were created by extracting and chronologically sorting COVID-19 phenotypes from the five categories: positive test, primary care diagnosis, hospitalisation, ICU admission and death. We focused the trajectory analysis on ICU admissions, rather than including all critical care treatments, as procedure dates were unavailable. Only the first date for each event category was considered, for multiple events recorded on the same day (e.g. positive test and hospitalisation) events were ordered by the hierarchy: positive test, primary care diagnosis, hospitalisation, ICU admission and death. Trajectory network plots were developed by extracting all pairwise transitions between events and calculating frequency of, and the median number of days between, all individuals experiencing this transition.

Kaplan-Meier (KM) plots were used to estimate 28-day COVID-19 mortality, stratified by worst healthcare presentation as a proxy for COVID-19 severity. Both trajectory and KM plots were stratified by pandemic waves, sex, age, ethnicity and socioeconomic deprivation. KM plots were also used to compare the cumulative event frequency of the five main COVID-19 phenotypes stratified by vaccination status.

Data were accessed through the NHS Digital Data Access Environment (DAE). Data cleaning, exploratory analysis, phenotype creation and cohort assembly was performed using Python (3.7) and Spark SQL (2.4.5) on Databricks Runtime 6.4 for Machine Learning. Analysis was performed in RStudio (Professional) Version 1.3.1093.1 - R Version 4.0.3. Summary statistics were created using tableOne (0.12.0). Figures were constructed using ggplot2 (3.3.3), VennDiagram (1.6.20), igraph (1.2.6), survival (3.2.7) and survminer (0.4.8) packages.

### Ethical and Regulatory Approvals

Ethical approval of the CVD-COVID-UK consortium has been described in detail previously^7^. Data access approval was granted to the CVD-COVID-UK consortium (under project proposal CCU013 High-throughput electronic health record phenotyping approaches) through the NHS Digital online Data Access Request Service^16^ (ref. DARS-NIC-381078-Y9C5K). NHS Digital data have been made available for research under the Control of Patient Information (COPI) notice which mandated the sharing of national electronic health records for COVID-19 research (more info: https://digital.nhs.uk/coronavirus/coronavirus-covid-19-response-information-governance-hub/control-of-patient-information-copi-notice). For further detail see supplementary methods.

## Results

Our cohort consisted of 56,609,049 individuals registered with a general practitioner in England and alive on 23rd January 2020, among whom we identified 8,825,738 COVID-19 events in 3,469,528 individuals, representing an infection rate of 6.1%, across the study period ending 31st March 2021 (Figure 1 & Supplementary Figure 1). The mean follow-up time from the first COVID-19 event was 124 days, with a minimum of 28 days. There were 3,114,784 individuals (90%) with a positive test, 2,363,507 (68%) with a COVID-19 diagnosis in primary care, 364,260 (11%) who were hospitalised and 60,184 (1.7%) who received critical care (encompassing ventilatory support and ICU admission).

Most individuals with COVID-19 event(s), (89%, n = 3,094,860) avoided hospitalisation or death related to COVID-19. Of those admitted to hospital, 54,026 (15%) received NIV, 38,072 (10%) were admitted to an ICU, 21,404 (6%) received IMV, 17,786 (4.9%) patients received both NIV and IMV and 549 received ECMO.

Demographic characteristics of patients are reported in table 1 with the entire dataset population (56.6 million) shown for comparison. A proportionately greater number of COVID-19 infections were observed in femalepatients (55%), patients of Asian ethnicity (13 %) and patients in the most deprived fifth (24%). The overall frequency of COVID-19 events follows the bimodal distribution of England’s first and second waves^9^ (Supplementary Figure 3 & 4).

**Table 1:** Sample characteristics of full population and individuals with COVID-19 by most severe COVID-19 phenotype. Values are numbers (percentages) unless otherwise specified. Deaths only, represents patients who did not have any other COVID-19 related healthcare presentation before dying with COVID-19 recorded on the death certificate. For comorbidities, [N] is the number of conditions included under each clinical specialties / organs system, count of individual identified conditions can be seen in supplementary table 3. Multimorbidity represents the binary sum across all 270 CALIBER phenotypes.

|  | Population | All COVID-19 Cases | Severity (worst healthcare presentation) |  |  |  |  | All COVID-19 Deaths |
| --- | --- | --- | --- | --- | --- | --- | --- | --- |
|  |  |  | Positive Test | Primary Care Diagnosis | Hospitalisation | Critical Care | Deaths only |  |
| n (%) | 56608985 | 3469528 (6.1) | 931126 (26.8) | 2163734 (62.4) | 304081 (8.8) | 60184 (1.7) | 10403 (0.3) | 140908 |
| COVID Deaths (%) | 140908 (0.2) | 140908 (4.1) | 9580 (1.0) | 19527 (0.9) | 75439 (24.8) | 25959 (43.1) | 10403 (100.0) | 140908 (100.0) |
| All Deaths (%) | 723925 (1.3) | 178721 (5.2) | 13654 (1.5) | 34006 (1.6) | 93944 (30.9) | 26714 (44.4) | 10403 (100.0) | 140298 (99.6) |
| Male (%) | 28031640 (49.5) | 1571566 (45.3) | 428342 (46.0) | 945768 (43.7) | 154210 (50.7) | 38385 (63.8) | 4861 (46.7) | 76167 (54.1) |
| Age (%) |  |  |  |  |  |  |  |  |
| Under 18 | 12154477 (21.5) | 385744 (11.1) | 114162 (12.3) | 266481 (12.3) | 4550 (1.5) | 549 (0.9) | 2 (0.0) | 57 (0.0) |
| 18 - 29 | 8861746 (15.7) | 708501 (20.4) | 227635 (24.4) | 467208 (21.6) | 12380 (4.1) | 1261 (2.1) | 17 (0.2) | 261 (0.2) |
| 30 - 49 | 15561629 (27.5) | 1131044 (32.6) | 331697 (35.6) | 748555 (34.6) | 41119 (13.5) | 9453 (15.7) | 220 (2.1) | 3164 (2.2) |
| 50 - 69 | 13265698 (23.4) | 824537 (23.8) | 200424 (21.5) | 516217 (23.9) | 78515 (25.8) | 28275 (47.0) | 1106 (10.6) | 22626 (16.1) |
| Over 70 | 6765435 (12.0) | 419702 (12.1) | 57208 (6.1) | 165273 (7.6) | 167517 (55.1) | 20646 (34.3) | 9058 (87.1) | 114800 (81.5) |
| Ethnic group (%) |  |  |  |  |  |  |  |  |
| White | 45300233 (80.0) | 2679541 (77.2) | 717592 (77.1) | 1658731 (76.7) | 249964 (82.2) | 43987 (73.1) | 9267 (89.1) | 123738 (87.8) |
| Asian or Asian British | 4892279 (8.6) | 448329 (12.9) | 109235 (11.7) | 299881 (13.9) | 29529 (9.7) | 9191 (15.3) | 493 (4.7) | 9286 (6.6) |
| Black or Black British | 2155780 (3.8) | 139171 (4.0) | 38733 (4.2) | 82924 (3.8) | 13406 (4.4) | 3800 (6.3) | 308 (3.0) | 4111 (2.9) |
| Chinese | 516056 (0.9) | 10412 (0.3) | 2924 (0.3) | 6271 (0.3) | 871 (0.3) | 319 (0.5) | 27 (0.3) | 333 (0.2) |
| Mixed | 1198711 (2.1) | 68536 (2.0) | 20794 (2.2) | 42912 (2.0) | 3796 (1.2) | 964 (1.6) | 70 (0.7) | 976 (0.7) |
| Other | 1249555 (2.2) | 73041 (2.1) | 21875 (2.3) | 44536 (2.1) | 5047 (1.7) | 1461 (2.4) | 122 (1.2) | 1472 (1.0) |
| Unknown | 1296371 (2.3) | 50498 (1.5) | 19973 (2.1) | 28479 (1.3) | 1468 (0.5) | 462 (0.8) | 116 (1.1) | 992 (0.7) |
| Social deprivation fifths (%) |  |  |  |  |  |  |  |  |
| 1 (most deprived) | 11702944 (20.7) | 832546 (24.0) | 212631 (22.8) | 521632 (24.1) | 79559 (26.2) | 16564 (27.5) | 2160 (20.8) | 32786 (23.3) |
| 5 (least deprived) | 10816336 (19.1) | 562664 (16.2) | 150872 (16.2) | 355623 (16.4) | 45842 (15.1) | 8275 (13.7) | 2052 (19.7) | 23743 (16.9) |
| Unknown | 53813 (0.1) | 2902 (0.1) | 700 (0.1) | 1876 (0.1) | 265 (0.1) | 39 (0.1) | 22 (0.2) | 127 (0.1) |
| Comorbidities (%) |  |  |  |  |  |  |  |  |
| High risk (Shielding List) | 4030761 (7.1) | 381497 (11.0) | 65551 (7.0) | 184741 (8.5) | 110632 (36.4) | 18713 (31.1) | 1860 (17.9) | 41446 (29.4) |
| Circulatory [35] | 10851482 (19.2) | 736898 (21.2) | 131253 (14.1) | 368466 (17.0) | 193261 (63.6) | 35716 (59.3) | 8202 (78.8) | 113441 (80.5) |
| Respiratory [16] | 9899255 (17.5) | 727167 (21.0) | 170862 (18.4) | 432552 (20.0) | 100218 (33.0) | 20419 (33.9) | 3116 (30.0) | 49350 (35.0) |
| Genitourinary [17] | 3882322 (6.9) | 336029 (9.7) | 65672 (7.1) | 174592 (8.1) | 79398 (26.1) | 12819 (21.3) | 3548 (34.1) | 46213 (32.8) |
| Digestive [25] | 5191652 (9.2) | 381476 (11.0) | 81658 (8.8) | 213462 (9.9) | 70698 (23.2) | 13329 (22.1) | 2329 (22.4) | 35966 (25.5) |
| Endocrine [7] | 7614040 (13.5) | 590182 (17.0) | 119649 (12.8) | 316538 (14.6) | 122759 (40.4) | 27104 (45.0) | 4132 (39.7) | 64079 (45.5) |
| Haematological/Immunological [18] | 663089 (1.2) | 56301 (1.6) | 11131 (1.2) | 29251 (1.4) | 12964 (4.3) | 2607 (4.3) | 348 (3.3) | 6448 (4.6) |
| Infectious Diseases [26] | 8607760 (15.2) | 697744 (20.1) | 182169 (19.6) | 380435 (17.6) | 113468 (37.3) | 16304 (27.1) | 5368 (51.6) | 64982 (46.1) |
| Neurological [12] | 1825348 (3.2) | 147025 (4.2) | 28156 (3.0) | 74165 (3.4) | 37157 (12.2) | 6146 (10.2) | 1401 (13.5) | 19706 (14.0) |
| Cancers [41] | 5847529 (10.3) | 409594 (11.8) | 118612 (12.7) | 225649 (10.4) | 54435 (17.9) | 9120 (15.2) | 1778 (17.1) | 28421 (20.2) |
| Benign Neoplasms [7] | 2731495 (4.8) | 196641 (5.7) | 39771 (4.3) | 112528 (5.2) | 36576 (12.0) | 6761 (11.2) | 1005 (9.7) | 17473 (12.4) |
| Mental Health [13] | 12184675 (21.5) | 895649 (25.8) | 204806 (22.0) | 531492 (24.6) | 130223 (42.8) | 22247 (37.0) | 6881 (66.1) | 71351 (50.6) |
| Musculoskeletal [19] | 4265188 (7.5) | 320870 (9.2) | 58939 (6.3) | 164354 (7.6) | 81789 (26.9) | 11663 (19.4) | 4125 (39.7) | 50164 (35.6) |
| Skin [9] | 1514761 (2.7) | 106835 (3.1) | 26516 (2.8) | 63272 (2.9) | 14107 (4.6) | 2454 (4.1) | 486 (4.7) | 7125 (5.1) |
| Eye [12] | 4740276 (8.4) | 357027 (10.3) | 56878 (6.1) | 159795 (7.4) | 115292 (37.9) | 20056 (33.3) | 5006 (48.1) | 71302 (50.6) |
| Ear [3] | 180196 (0.3) | 14502 (0.4) | 2234 (0.2) | 6672 (0.3) | 4834 (1.6) | 550 (0.9) | 212 (2.0) | 2896 (2.1) |
| Perinatal [9] | 1580818 (2.8) | 61883 (1.8) | 17428 (1.9) | 41319 (1.9) | 2577 (0.8) | 506 (0.8) | 53 (0.5) | 768 (0.5) |
| Multimorbidity (median [IQR]) | 1.00 [0, 3] | 1.00 [0, 4] | 1.00 [0, 3] | 1.00 [0, 3] | 6.00 [3, 10] | 5.00 [2, 8] | 7.00 [5, 11] | 8.00 [5, 11] |

### COVID-19 mortality

140,908 individuals died, representing a mortality rate of 4%. Of these deaths, the majority occurred in patients who were hospitalised (72%, n=101,398), however 39,511 (28%) died without having ever been admitted to hospital. Notably we identified 10,403 unheralded COVID-19 deaths - individuals who died with COVID-19 as a recorded cause, but for whom no other COVID-19 phenotypes, such as positive tests or primary care diagnosis, were identified. These individuals were overwhelmingly elderly (87% over 70), white (89%) and multimorbid (median 7 conditions) (Table 1).

13,083 individuals died within 28-days of a COVID-19 event without a confirmed or suspected COVID-19 diagnosis listed on the death certificate. Compared with those who died with COVID-19 as a recorded cause of death, these individuals were less likely to have a positive COVID-19 test, and had fewer admissions and ventilatory treatments. Their most frequent primary causes of death were: unspecified dementia (6.8%), pneumonia (5.4%), and cancer of bronchus and lung (5%) (supplementary table 4 & 5).

Comparing demographics for those dying of COVID-19 against all cases we can see that individuals in whom infection was fatal, more were male (54%), elderly (82% over 70), white (88%) and deprived (23% in most deprived fifth). Multimorbidity was significantly higher in these patients, with a median of 8 conditions, compared to 1 in all COVID cases, 6 in those hospitalised and 5 in those receiving critical care. Amongst the 4,071,794 individuals classified as “high risk”, 381,497 (9.4%) contracted COVID-19 and 41,446 died, giving a mortality rate of 11%.

### Mortality in hospitalised patients

Patients receiving critical care outside ICU had the highest mortality (47%, n=10,497), compared with those admitted to ICU (41%, n=15,462) and those admitted to hospital but not receiving critical care (25%, n=75,439). Table 2 compares characteristics for individuals who were admitted to hospital, admitted to ICU, received critical care outside ICU and who died during waves 1 and 2. Mortality amongst all hospitalised patients decreased from 32.6% to 26.4%. The overall proportion of hospitalised patients receiving critical care and admitted to ICU did not significantly change, however an increase in numbers receiving NIV (13.9% to 15.4%), and a corresponding decrease in IMV (7.1% to 5.4%), coupled with a small increase in numbers receiving critical care outside ICU (5.5% to 6.4%) was observed in wave 2. Despite these changes, the mortality of those admitted to ICU did not significantly differ between waves (40.4 vs 41.1% p = 0.226), in contrast to the improvement in mortality for inpatients not receiving critical care (30.4% to 23.1%) and those receiving critical care outside ICU (50.7% to 46.3%).

**Table 2:**
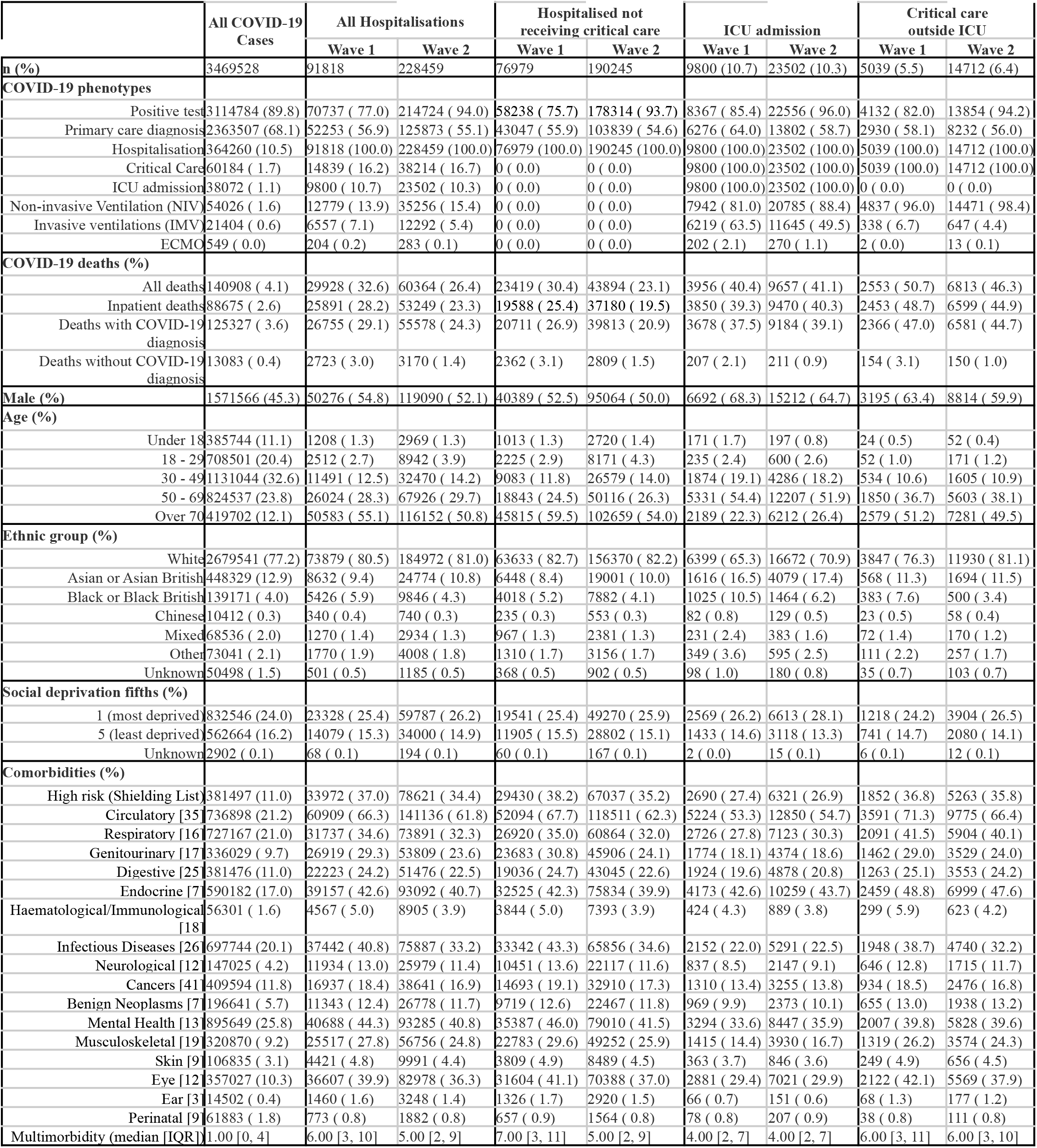
Comparison between pandemic wave 1 and 2 of people with COVID-19 who were hospitalised, in critical care or died. Values are numbers (percentages) unless otherwise specified. For comorbidities, [N] is the number of conditions included under each clinical specialties / organs system, count of individual identified conditions can be seen in supplementary table 3.

### Trajectories across pandemic waves

Kaplan-Meier survival analysis and trajectory analysis provide methods to study temporal changes in disease progression between pandemic waves. Kaplan-Meier curves for 28 day mortality corroborate the finding that overall mortality does not change for those admitted to ICU whilst revealing a prolongation of survival time reflected by the reduced slope of the KM curve in the second wave (Figure 3). This change in survival time is consistent across those hospitalised and not admitted to ICU.

**Figure 2:**
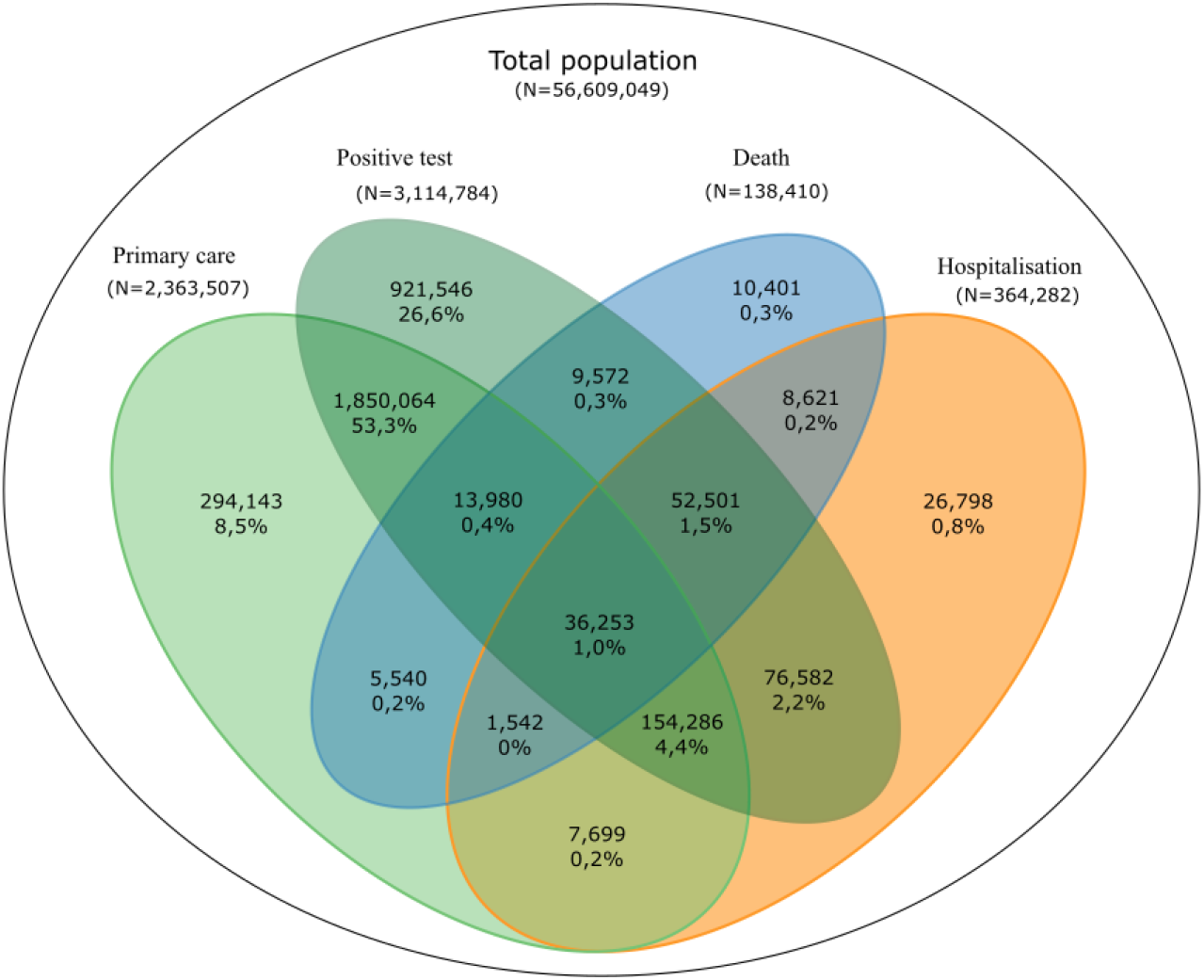
Venn Diagram of individuals with one or more of the 10 COVID-19 severity phenotypes across four groups of data sources. COVID-19 testing (SGSS), primary care (GDPPR), secondary care (HES APC, SUS, HES CC, and CHESS) and deaths (ONS deaths registry). Numbers indicate unique individuals identified in 1, 2, 3 or 4 sources from N=3,469,528 unique individuals with COVID-19.

**Figure 3:**
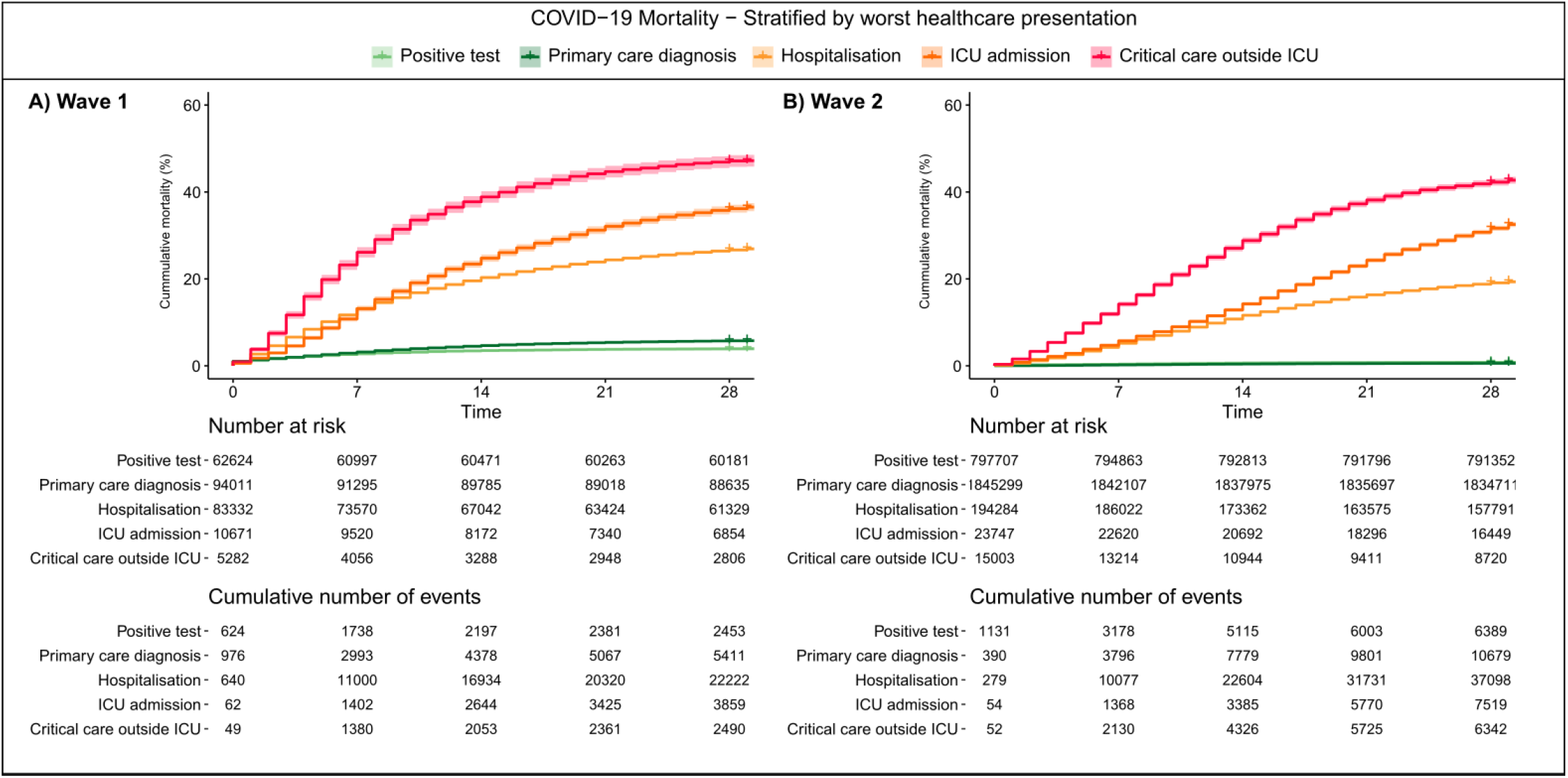
Cumulative Covid-19 mortality events stratified by most severe COVID-19 phenotype. Plots are stratified by A) wave 1 and B) wave 2. Shaded areas represent 95% confidence intervals.

Trajectory analysis provides further insight into the temporal progression of phenotypes and mirrors our other findings by demonstrating an increase in median days between a positive test, primary care diagnosis or hospitalisation and death of 4, 5 & 3 days respectively for the second wave as compared to the first (Figure 4). Trajectories and KM curves stratified by age, sex, ethnicity and deprivation are available in the supplement.

**Figure 4:**
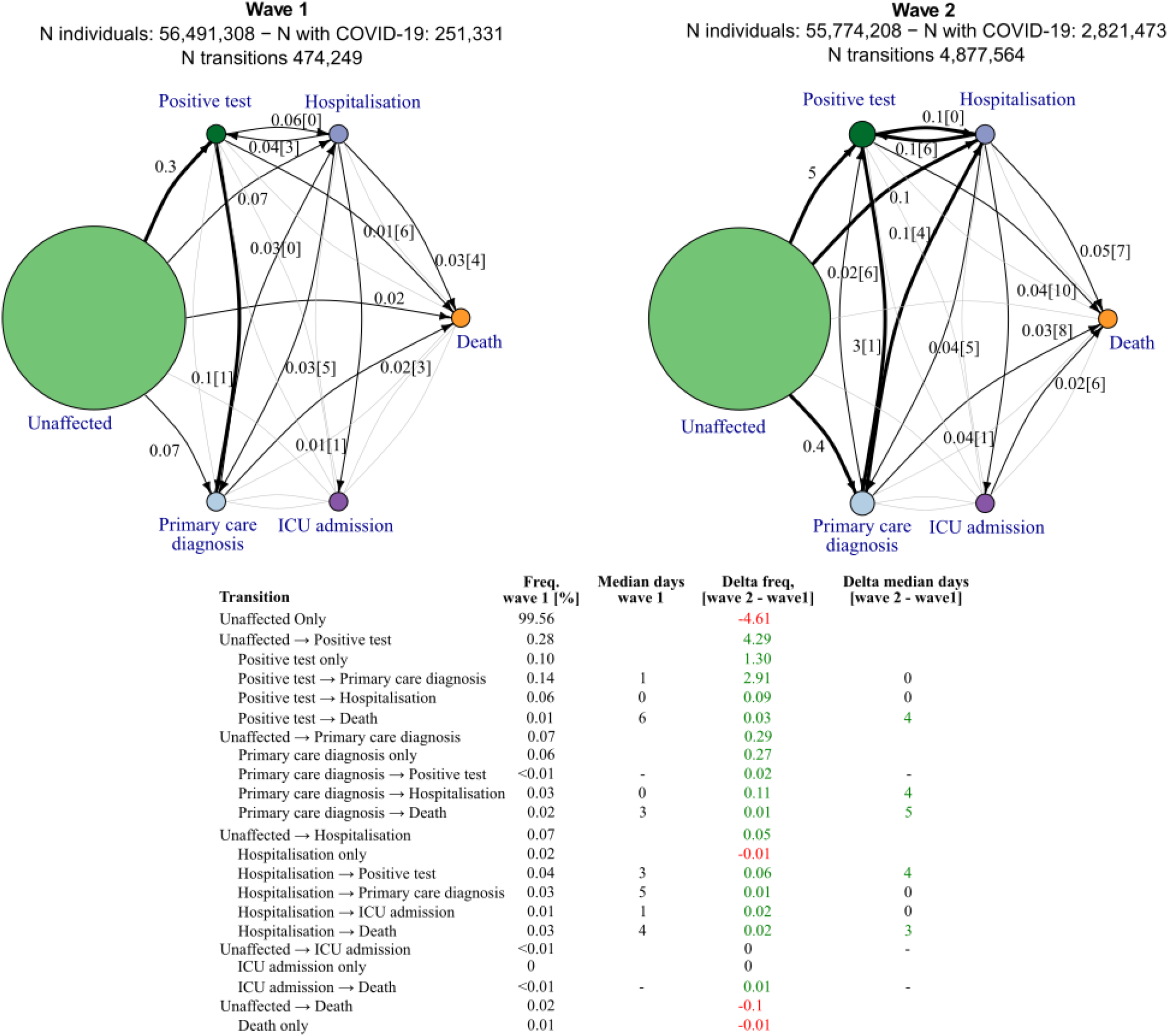
COVID-19 trajectory networks, showing percentage of individuals transitioning and the median number of days passing between severity phenotypes in pandemic wave 1 and 2. The size of the circles represent the number of individuals with that event relative to the total study population size of 56.6 million. Numbers on arrows are the percentage of individuals with the given transition (relative to N individuals in the given wave) and in square brackets median days between events across all individuals with that transition. Median days between unaffected and other severity phenotypes are not shown as they are not directly comparable between waves, due to difference in length of the two periods. Thick arrows represent transitions occurring in ≥ 0.1%. Thin black arrows represent transitions occurring in ≥ 0.01%. Any transitions occurring in fewer than 0.01% are not shown. All included individuals were all alive and had no previous COVID-19 events recorded before the start date of the specified waves.

**Figure 5:**
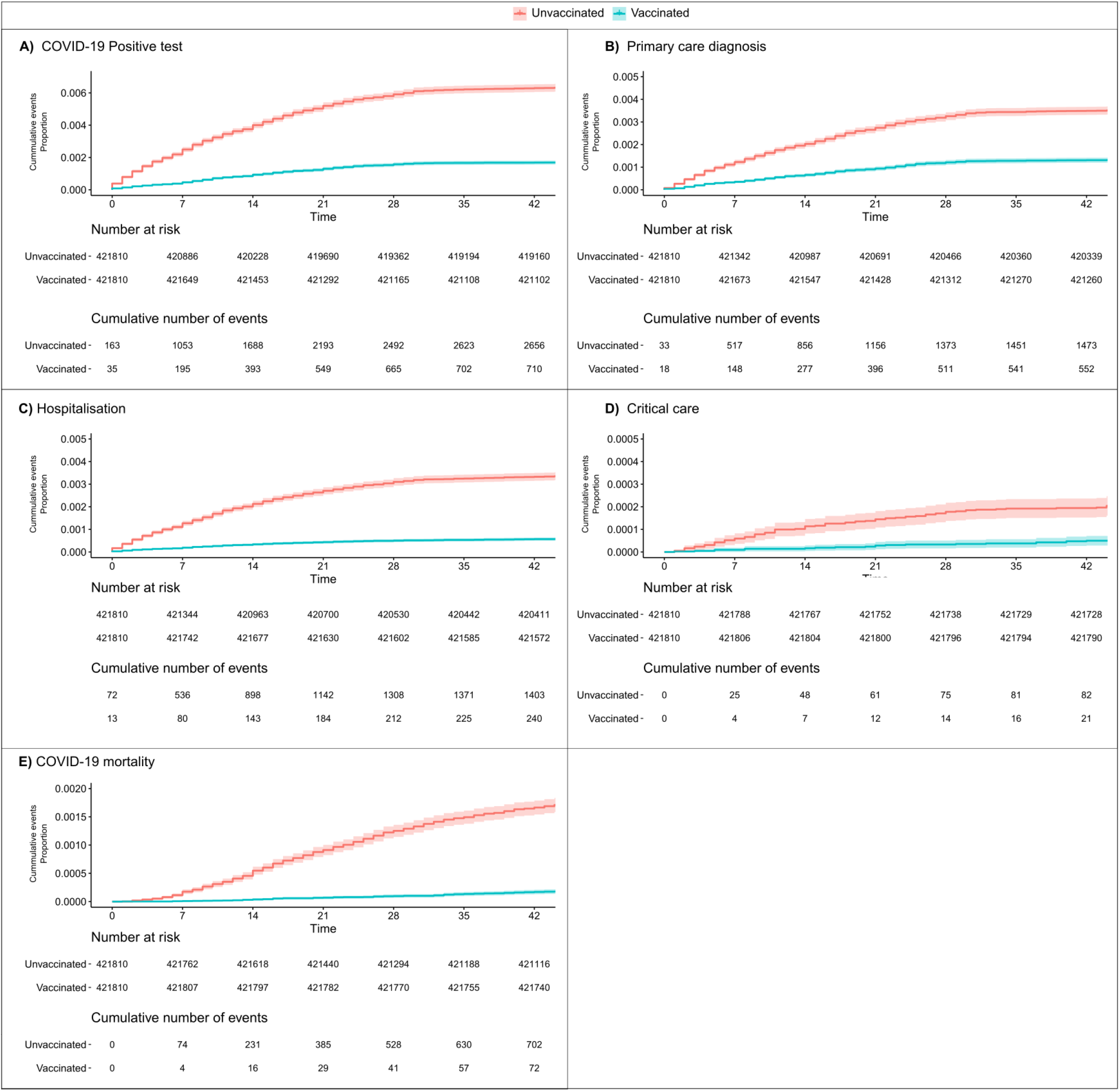
Kaplan-Meier plots of cumulative COVID-19 events. Analysis all run from 1st of February 2021 and displays cumulative events of A) Covid-19 positive tests, and B) primary care diagnosis, C) hospitalisation, D) critical care and E) COVID-19 mortality in matched groups of fully vaccinated (two dosages a minimum 14 days prior to 01.02.2021) vs unvaccinated (no doses given before or through follow up). Matching was done on gender, five year age groups and ethnicity.

### Recording patterns across sources

No single source captured all COVID-19 cases (Figure 2). Approximately half of individuals with a positive test also received a primary care diagnosis, while 939,512 (27%) had a positive test but no other record. 12.2% with a primary care record had no other evidence of COVID-19, as did 7.1% with a secondary care record, and 10,403 COVID-19 cases were identified exclusively from mortality data with no prior COVID-19 events (Figure 2). A small number of individuals were identified only from PHE hospital surveillance data (CHESS, 2,720 individuals, 0.8% of all hospitalisations). See Supplementary Figure 2, for further details on data source overlap.

## Discussion

In this study we provide a comprehensive examination of COVID-19 disease recording patterns, severity and patient trajectories, across pandemic waves, using linked EHR from 56 million people in England. In contrast with existing research, which has been undertaken in clinical populations, or has solely used administrative data, our study utilised eight complementary datasets spanning healthcare settings on a national level and capturing a diverse set of disease exposures and outcomes. We defined and evaluated ten COVID-19 phenotypes, associated with five severity categories, and explored disease trajectories and mortality between pandemic waves. COVID-19 testing and treatment has been carried out through a number of different routes which have evolved during the pandemic and our findings illustrate the importance of linking multiple data sources to maximise event ascertainment, fully capturing the spectrum of potential health outcomes and identifying patient transitions through the healthcare system. The phenotypes presented here have already enabled other analyses with highly-relevant public health policy implications such as assessing the association of COVID-19 vaccines ChAdOx1 and BNT162b2 with major venous, arterial, and thrombocytopenic events^17^, and evaluating the use of antithrombotic medication on COVID-19 outcomes^18^ and studying the incidence of vascular diseases post COVID-19 infection^19^.

Our study expands the literature in several key ways. To our knowledge, it is the largest study in terms of sample size and data fidelity to create and evaluate computable COVID-19 phenotypes by leveraging multiple sources of linked data on a national level spanning electronic health records, administrative claims, disease audits and national registers. The use of multiple sources enabled the ascertainment of events which would have previously been missed (for example, 9% of infected individuals died without COVID-19 being listed as a cause of death) or incorrectly aggregated (for example, patients that received ventilation outside of ICU had the highest mortality). The framework for defining disease severity across multiple settings can be adapted and applied in other countries with similar national/regional EHR sources (e.g. Denmark, Korea, Canada) and be used to monitor and assess pandemic impacts.

### Phenotype validation

We utilised the CALIBER phenotyping approach^10^ and performed multiple layers of validation. We internally validated our COVID-19 phenotypes through demonstrating cross-EHR source concordance. External validation was provided through our analysis of patient demographics and comorbidities which is consistent with widely reported associations between sex, age, ethnicity and deprivation and outcomes for patients in primary care^3^, secondary care^20^ and ICU^21^. Additionally, our ascertained infection rate of 6.18%, was comparable with official estimates from PHE^9^ and the National Real-time Assessment of Community Transmission (REACT) study^22^. Finally, comparison of 28 day cumulative event frequency of the five main phenotype categories between those fully vaccinated and unvaccinated controls reproduced the known and expected protective effects of vaccination^2^.

### Ascertaining events across multiple EHR sources

A key challenge of using multiple data sources is the harmonisation of information across each source in the absence of a gold standard as each dataset contained information at different levels of resolution and reflected variations in healthcare delivery across the duration of the pandemic. For example, when creating phenotypes for ventilatory support, we exploited linkage across multiple EHR sources (HES APC, SUS, HES CC, and CHESS), alongside OPCS-4 procedure codes for ventilatory support modalities. This approach allowed us to identify 21,558 individuals who received NIV outside of ICU, representing 40% of all patients treated with NIV, and showed that patients receiving critical care outside ICU had the highest mortality. These important findings are consistent with COVID-19 overwhelming pre-existing critical care capacity and necessitating expansion of services to new areas, such as operating theatres and recovery wards^23^. Furthermore they illustrate the value of linkage and our rigorous phenotypes for maximising data capture, particularly in identifying groups that may otherwise ‘fall through the cracks’ between datasets, as in this case.

### Disease trajectories

The increased median duration between COVID-19 phenotypes observed in wave 2, when compared to wave 1, has several potential explanations including the increased availability of testing, leading to individuals being identified earlier in their infection, and changes in inpatient management, such as the widespread adoption of dexamethasone following the results of the RECOVERY trial^24^.

Stratification of trajectories by demographics revealed patterns including a decreased number of days from positive test and primary care diagnosis to death in the most deprived and non-white ethnicities. This may suggest these groups may be accessing healthcare later in their disease progression, for reasons which are likely multifactorial, but related to existing socioeconomic health inequalities exacerbated by the pandemic^25^.

### Implications

The COVID-19 phenotypes and trajectory analysis outlined here produce a reproducible, extensible and repurposable means to generate national-scale data to support critical policy decision making. By modelling patient trajectories as a series of interactions within the healthcare system, and linking these to demographic and outcome data, we provide a means to identify and prioritise care pathways associated with adverse outcomes and highlight healthcare system ‘touch points’ which may act as tangible targets for intervention - for example access to testing for the most deprived and non-white ethnicities. Beyond the pandemic we believe that trajectory analysis has the potential to transform analysis of complex conditions and multimorbidity through unlocking the power of linked data to disentangle the progression of individuals through the healthcare system and disease states over time.

In sharing fully reproducible analytical code and phenotypes we envisage that this work will facilitate other researchers to produce high quality and consistent outputs across a diverse range of topics. Linkage to additional datasets, as illustrated by vaccination data, allows extension to address new research questions, such as the emergence of novel variants, or assessing the efficacy of booster/third primary vaccine doses on both patient and healthcare system level outcomes. Thus we provide a framework by which real world healthcare data can be used to address questions of critical policy relevance and positively impact the care and safety of the nation.

### Strengths and limitations

A key strength of this work using national-scale data is that by definition it is representative of the general population across all age groups, ethnicities, deprivation levels and demographic characteristics. To our knowledge, this is the largest population-wide research study of COVID-19 phenotypes which includes: a) multiple healthcare settings through data linkage at a population level, b) detailed identification of specific ventilatory treatments, c) classification of COVID-19 related deaths, and d) exploration of transitions between COVID-19 events. Using multiple EHR sources spanning different healthcare settings, maximised infection ascertainment and reduced the effects of variable testing and data recording patterns (especially during the first wave).

As the focus of this work was to create COVID-19 related phenotypes, and describe the characteristics of individuals experiencing them, we have not conducted multivariable regression analyses to control for confounders. The findings presented are therefore not associative statements and should not be interpreted as causal relationships. However by sharing reproducible phenotype definitions we hope to facilitate further work to address the questions raised in this and other COVID-19 studies exploring national level data, as exemplified by recent research^17–19^.

Whilst our definitions of the pandemic waves differ from others, we believe using non-contiguous dates enabled a balanced comparison across periods of heightened strain than including the period of low cases over the summer in the first wave. The recording of dates in EHRs will not always be fully accurate. We sought to mitigate inaccuracies by reporting the median number of days, and through only reporting time differences between transitions happening in more than 0.01% of all transitions. Finally, data granularity was limited: for example we could not delineate whether a patient received NIV followed by IMV, represented an escalation of ventilatory support, or IMV followed by NIV, and therefore focused on ICU admissions where accurate start dates were available.

## Conclusion

Exploiting linkage across electronic health records on a national-scale allows us to highlight the healthcare trajectories of individuals with COVID-19, revealing who has been affected, and how.

Defining new phenotypes empowers analysts to look beyond binary outcomes such as mortality, to significant interim events, such as ventilatory treatments and ICU admission, and enables characterisation of an individuals’ progression through these states. Furthermore, trajectory analysis provides a method to link traditionally disaggregated datasets to provide insight into behaviour on a national-scale and enables unlocking insights from populations that may ‘fall through the cracks’ of other analysis methods, for example those dying outside of hospital, or receiving critical care outside of ICU.

As demonstrated for vaccination efficacy, this work provides an adaptable framework that may be rapidly repurposed to answer questions of critical clinical and policy relevance; the emergence of a new variant, the need for booster doses in the context of waning immunity or simply to maximise the value of existing healthcare data in understanding individual’s progression through complex chronic diseases.

## Supporting information

STROBE / RECORD statement

## Data Availability

Data access approval was granted to the CVD-COVID-UK consortium (under project proposal CCU013 High-throughput electronic health record phenotyping approaches) through the NHS Digital online Data Access Request Service (ref. DARS-NIC-381078-Y9C5K). Requests for data access should be made to NHS Digital.

https://digital.nhs.uk/services/data-access-request-service-dars

https://web.www.healthdatagateway.org/dataset/7e5f0247-f033-4f98-aed3-3d7422b9dc6d

## Acknowledgements

This work is carried out with the support of the BHF Data Science Centre led by HDR UK (BHF Grant no. SP/19/3/34678). This work uses data provided by patients and collected by the NHS as part of their care and support. We would like to acknowledge all data providers who make anonymised data available for research.

The views expressed are those of the authors and not necessarily those of the organisations listed. The funders of this work played no role in the collection, analysis, or interpretation of data; in the writing of the report; or in the decision to submit the article for publication.

Competing interests: All authors have completed the ICMJE uniform disclosure form at www.icmje.org/coi_disclosure.pdf and declare: support from the funders listed above; no financial relationships with any organisations that might have an interest in the submitted work in the previous three years; no other relationships or activities that could appear to have influenced the submitted work. SH works as a data scientist and data curator for NHS Digital, which holds and processes the data.

Data sharing: The authors and colleagues across the CVD-COVID-UK consortium have invested considerable time and energy in developing the data resource described here and are keen to ensure that it is used widely to maximise its value. For inquiries about data access, please see www.healthdatagateway.org/dataset/7e5f0247-f033-4f98-aed3-3d7422b9dc6d or.

The three lead authors, JT, CT and SD (the manuscript’s guarantors), affirm that the manuscript is an honest, accurate, and transparent account of the resource and analyses being described and that no important aspects have been omitted.

Dissemination to participants and related patient and public communities: Results will be disseminated through the British Heart Foundation (BHF) Data Science Centre and CVD-COVID-UK webpages on the Health Data Research UK website, BHF communication channels, the BHF Data Science Centre’s lay members panel, and NHS Digital communications channels.

The study team would like to thank the BHF Data Science Centre’s lay members panel for their input and NHS DAE output checkers Lisa Grat and James Walker.

## Funding

The British Heart Foundation Data Science Centre (grant No SP/19/3/34678, awarded to Health Data Research (HDR) UK) funded co-development (with NHS Digital) of the trusted research environment, provision of linked datasets, data access, user software licences, computational usage, and data management and wrangling support, with additional contributions from the HDR UK data and connectivity component of the UK governments’ chief scientific adviser’s national core studies programme to coordinate national covid-19 priority research. Consortium partner organisations funded the time of contributing data analysts, biostatisticians, epidemiologists, and clinicians.

This work was funded by the Longitudinal Health and Wellbeing COVID-19 National Core Study, which was established by the UK Chief Scientific Officer in October 2020 and funded by UK Research and Innovation (grant references MC_PC_20030 and MC_PC_20059).

This work was supported by Health Data Research UK, which receives its funding from HDR UK Ltd (HDR-9006) funded by the UK Medical Research Council, Engineering and Physical Sciences Research Council, Economic and Social Research Council, Department of Health and Social Care (England), Chief Scientist Office of the Scottish Government Health and Social Care Directorates, Health and Social Care Research and Development Division (Welsh Government), Public Health Agency (Northern Ireland), British Heart Foundation (BHF) and the Wellcome Trust.

AA is supported by Health Data Research UK (HDR-9006), which receives its funding from the UK Medical Research Council (MRC), Engineering and Physical Sciences Research Council (EPSRC), Economic and Social Research Council (ESRC), Department of Health and Social Care (England), Chief Scientist Office of the Scottish Government Health and Social Care Directorates, Health and Social Care Research and Development Division (Welsh government), Public Health Agency (Northern Ireland), British Heart Foundation (BHF), and Wellcome Trust; and Administrative Data Research UK, which is funded by the ESRC (grant ES/S007393/1).

AB is supported by research funding from the National Institute for Health Research (NIHR), British Medical Association, Astra-Zeneca, and UK Research and Innovation.

AB, AW, HH, and SD are part of the BigData@Heart Consortium, funded by the Innovative Medicines Initiative-2 Joint Undertaking under grant agreement No 116074.

AW and SI are supported by the BHF-Turing Cardiovascular Data Science Award (BCDSA\100005) and by core funding from UK MRC (MR/L003120/1), BHF (RG/13/13/30194; RG/18/13/33946), and NIHR Cambridge Biomedical Research Centre (BRC-1215-20014).

JAC and JS are supported by the Health Data Research (HDR) UK South West Better Care Partnership and the NIHR Bristol Biomedical Research Centre at University Hospitals Bristol, and Weston NHS Foundation Trust and the University of Bristol.

SD, HH are supported by HDR UK London, which receives its funding from HDR UK funded by the UK MRC, EPSRC, ESRC, Department of Health and Social Care (England), Chief Scientist Office of the Scottish Government Health and Social Care Directorates, Health and Social Care Research and Development Division (Welsh government), Public Health Agency (Northern Ireland), BHF, and Wellcome Trust; HH and SD are supported by the NIHR Biomedical Research Centre at University College London Hospital NHS Trust. SD is supported by an Alan Turing Fellowship (EP/N510129/1). HH is a NIHR Senior Investigator. SD and HH are supported by the BHF Accelerator Award AA/18/6/24223.

CT is supported by a UCL UKRI Centre for Doctoral Training in AI-enabled Healthcare studentship (EP/S021612/1), MRC Clinical Top-Up and a studentship from the NIHR Biomedical Research Centre at University College London Hospital NHS Trust.

WW is supported by a Scottish senior clinical fellowship, CSO (SCAF/17/01).

## Notes

### Competing Interest Statement

The authors have declared no competing interest.

### Clinical Protocols

https://github.com/BHFDSC/CCU013_01_ENG-COVID-19_event_phenotyping/tree/main/protocol

### Author Declarations

Data access approval was granted to the CVD-COVID-UK consortium (under project proposal CCU013 High-throughput electronic health record phenotyping approaches) through the NHS Digital online Data Access Request Service (ref. DARS-NIC-381078-Y9C5K). NHS Digital data have been made available for research under the Control of Patient Information (COPI) notice which mandated the sharing of national electronic health records for COVID-19 research (more info: https://digital.nhs.uk/coronavirus/coronavirus-covid-19-response-information-governance-hub/control-of-patient-information-copi-notice). For further detail see supplementary methods.

