## Supplement for "Understanding COVID-19 trajectories from a nationwide linked electronic health record cohort of 56 million people: phenotypes, severity, waves & vaccination"

### Supplementary Material

#### Supplementary methods

##### COVID-19 event Phenotyping

COVID-19 positive tests were defined as a positive result from national testing data (SGSS), encompassing tests from NHS hospitals for those with a clinical need and healthcare workers (known as ‘Pillar 1’) and swab testing from the wider population (known as ‘Pillar 2’). COVID-19 primary care diagnoses were identified from primary care (GDPPR) using SNOMED-CT terms. COVID-19 hospital admissions were defined as any hospital admission recorded in CHESS (COVID-19 specific hospitalisations data) or admissions with a COVID-19 diagnosis in HES APC or SUS as primary or other listed cause of hospitalisation.

Provision of critical care was ascertained from several sources: a) patients with an ICU admission recorded in CHESS or patients with an ICU admission (defined as an entry within HES Adult Critical Care) within the same admission as HES APC, b) patients receiving NIV, identified using the OPCS4 code E85.2 (Non-invasive ventilation NEC) or E85.6 (Continuous positive airway pressure), from SUS or HES APC, or a positive number of days receiving basic respiratory support in HES Adult Critical Care, or ‘*high flow nasal oxygen’* or ‘*non invasive mechanical ventilation’* recorded in CHESS, c) patients receiving Intermittent Mandatory Ventilation (IMV) were identified using OPCS4 code E85.1 (Invasive ventilation) or X56 (Intubation of trachea), in SUS or HES APC, or a non-zero entry for days receiving advanced respiratory support from HES Adult Critical Care, or ‘*invasive mechanical ventilation’* recorded in CHESS, d) patients receiving Extracorporeal Membrane Oxygenation (ECMO) identified using OPCS4 X58.1 (Extracorporeal membrane oxygenation) in SUS or HES APC or ‘*respiratory support ECMO’* recorded in CHESS.

Fatal COVID-19 events were identified from ONS Civil Registration of Deaths and secondary care (HES APC, SUS) and defined as: a) a suspected or confirmed COVID-19 diagnosis ICD-10 term present in any position on the death certificate, b) death within 28-days of the first recorded COVID-19 event (positive test, diagnosis or admission), irrespective of the cause of death recorded on the death certificate, or c) a COVID-19 hospital admission with a discharge method or destination denoting death, irrespective of cause and duration after the index event. See supplementary table 2 for the specific codes used.

Phenotype definitions were reviewed by clinicians, health data scientists and epidemiologists and validated by quantifying cross-EHR source concordance and checking consistency of findings with established risk factors from the literature, in keeping with the CALIBER approach[10].

##### Ethical and Regulatory Approvals

Data access approval was granted to the CVD-COVID-UK consortium (under project proposal CCU013 High-throughput electronic health record phenotyping approaches) through the NHS Digital online Data Access Request Service [16] (ref. DARS-NIC-381078-Y9C5K). For full detail see supplementary methods. The BHF Data Science Centre approvals and oversight board deemed that this project's work fell within the scope of the consortium’s ethical and regulatory approvals. Analyses were conducted by three approved researchers (JHT, CT, SD) via secure remote access to the TRE. Only summarised, aggregate results were exported, following manual review by the NHS Digital ‘safe outputs’ escrow service, to ensure no output placed in the public domain contains information that may be used to identify an individual [7]. The North East-Newcastle and North Tyneside 2 research ethics committee provided ethical approval for the CVD-COVID-UK research programme (REC No 20/NE/0161).

#### Supplementary Figures

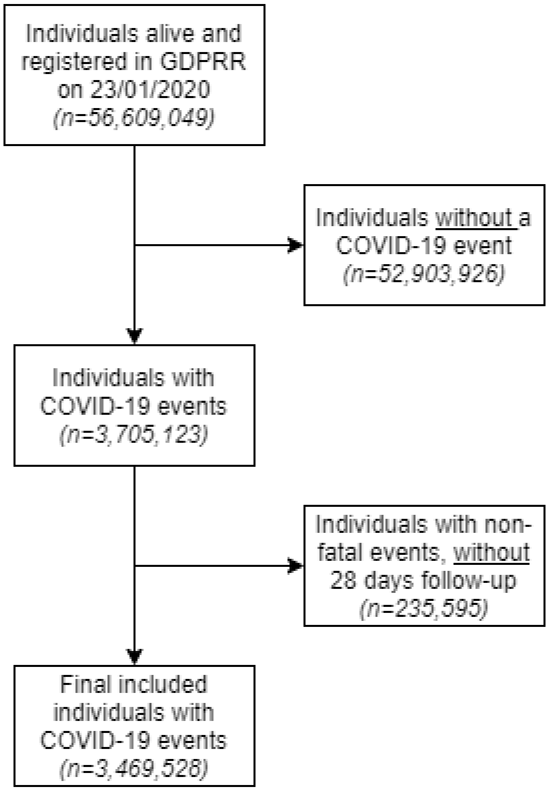


**Supplement figure 1: Flowchart of cohort design showing the number of records/individuals**
Excluded individuals at different stages and the identification of cases and the final study population.

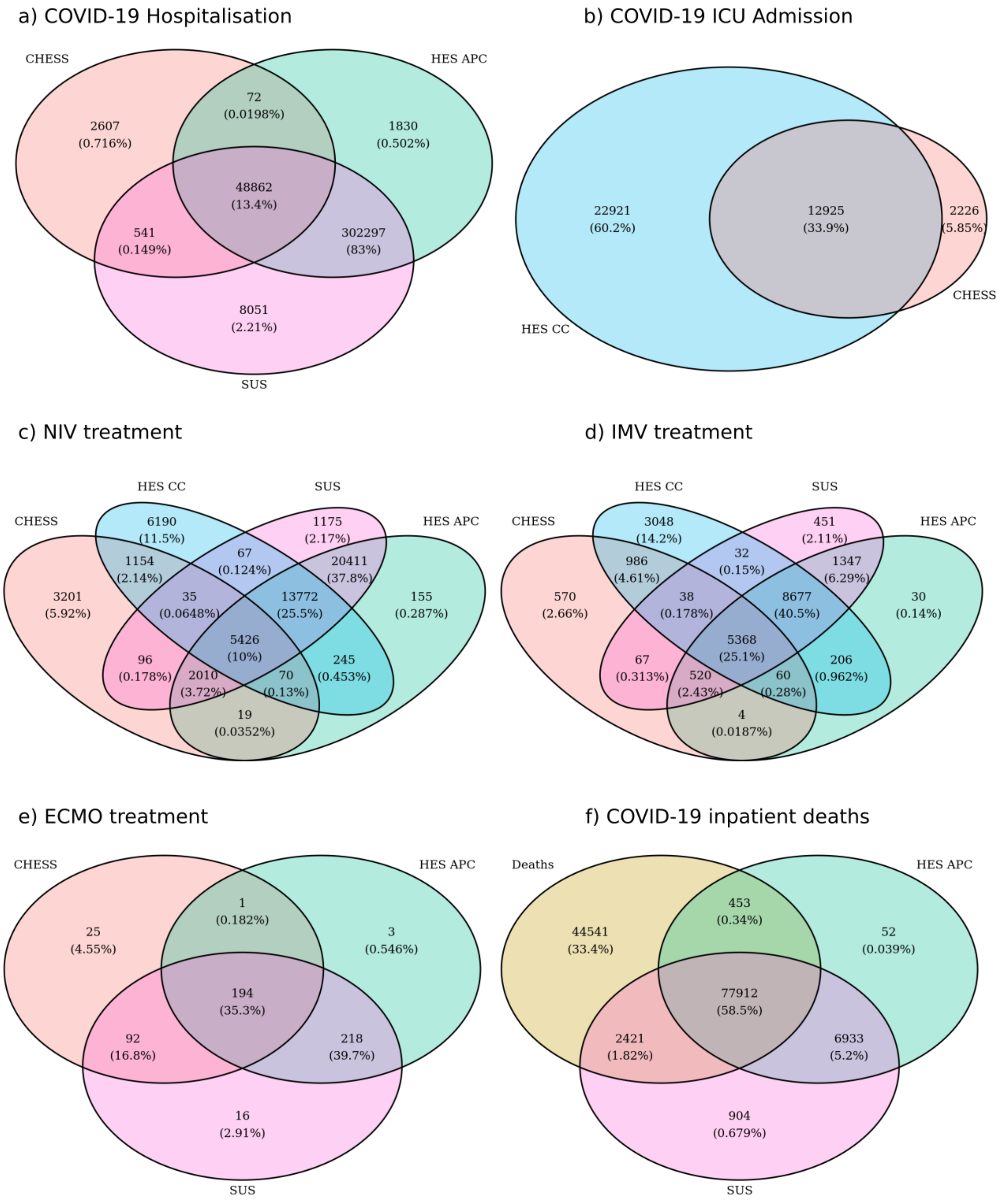


**Supplement figure 2: Venn Diagrams illustrating the numbers of individuals experiencing each COVID-19 event.**
Positive tests, primary care diagnoses and deaths with COVID-19 diagnosis, or within 28 days of a positive test, are not shown as these are derived from a single data source.

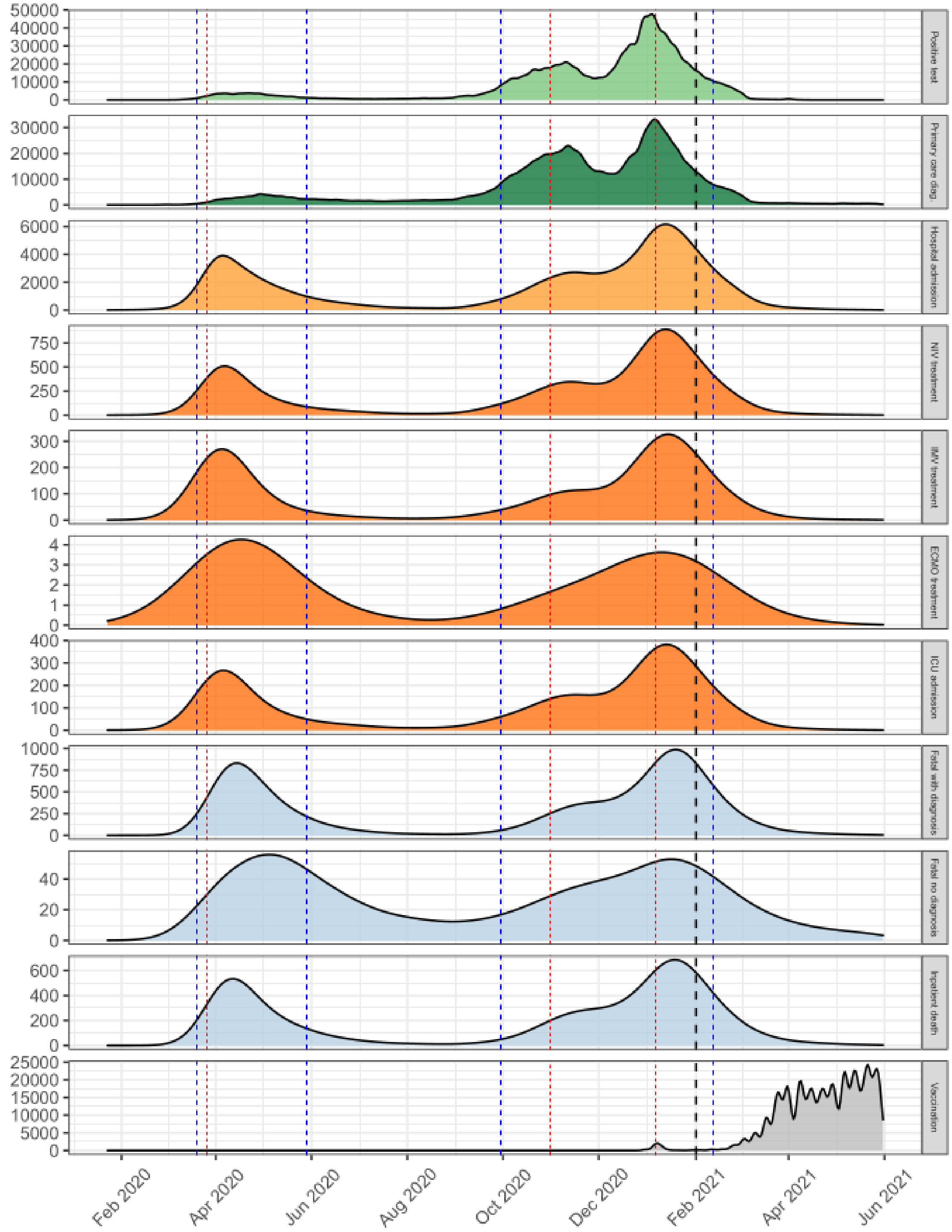


**Supplement figure 3: Timeline of COVID-19 event phenotypes.**
Kernel density estimation plot showing unique events per individual per date, a person may have multiple events of the same type at different dates. Vaccination shows the date of the second dosage. Red vertical lines indicate the official English lockdown dates (26.03.2020, 31.10.2020 & 06.01.2021). Blue vertical lines indicate our study definition of wave 1 (20.03.2020 - 29.05.2020) and wave 2 (30.09.2020 - 12.02.2021). Black vertical line indicates the date used to explore the effects of vaccination on COVID-19 phenotypes.

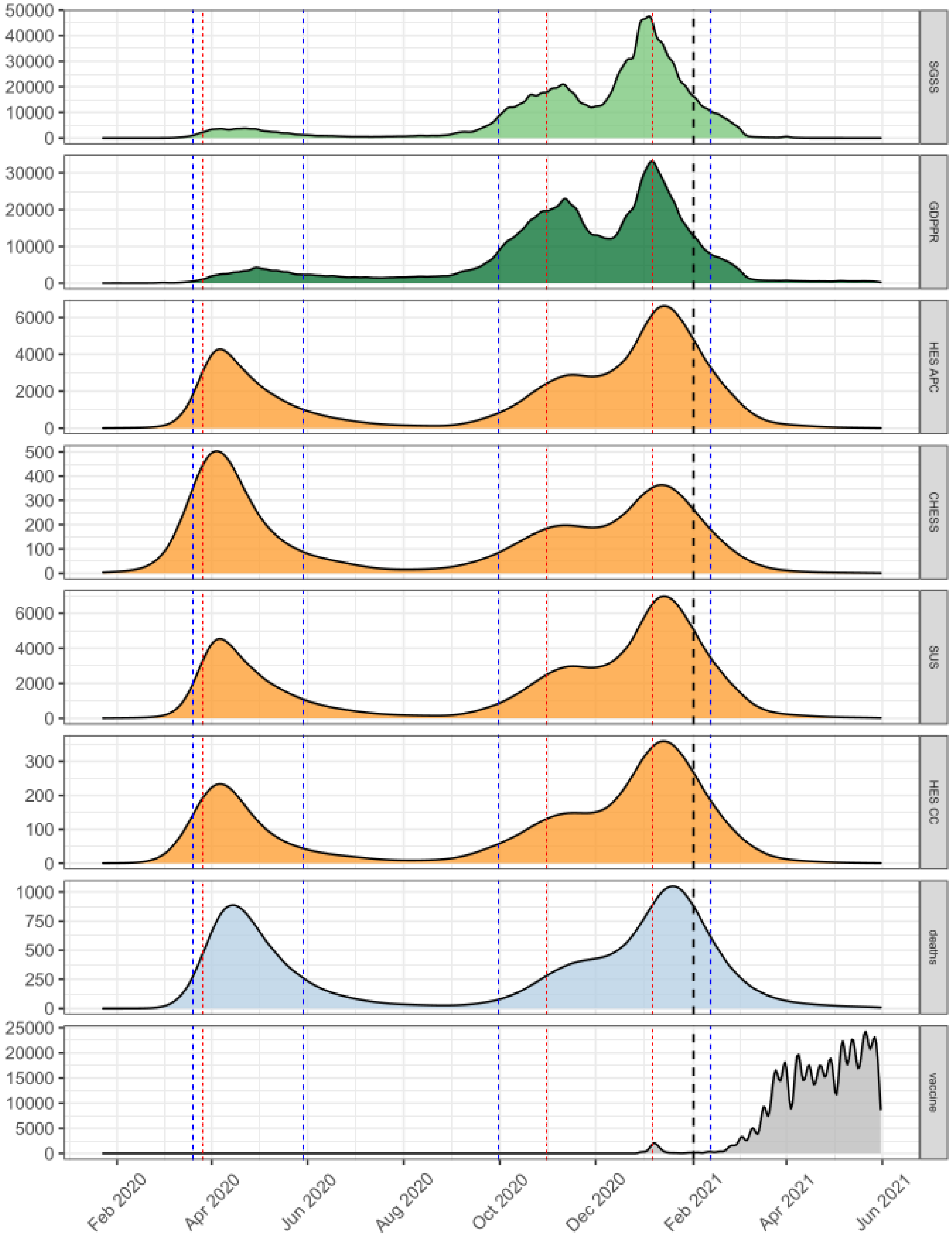


**Supplement figure 4: Timeline plots showing COVID-19 events, stratified by data source.**
A person may have multiple events from the same source at different dates. Vaccination shows the date of the second dosage. Red vertical lines indicate the official English lockdown dates (26.03.2020, 31.10.2020 & 06.01.2021). Blue vertical lines indicate our study definition of wave 1 (20.03.2020 - 29.05.2020) and wave 2 (30.09.2020 - 12.02.2021). Black vertical line indicates the date used to explore the effects of vaccination on COVID-19 phenotypes.

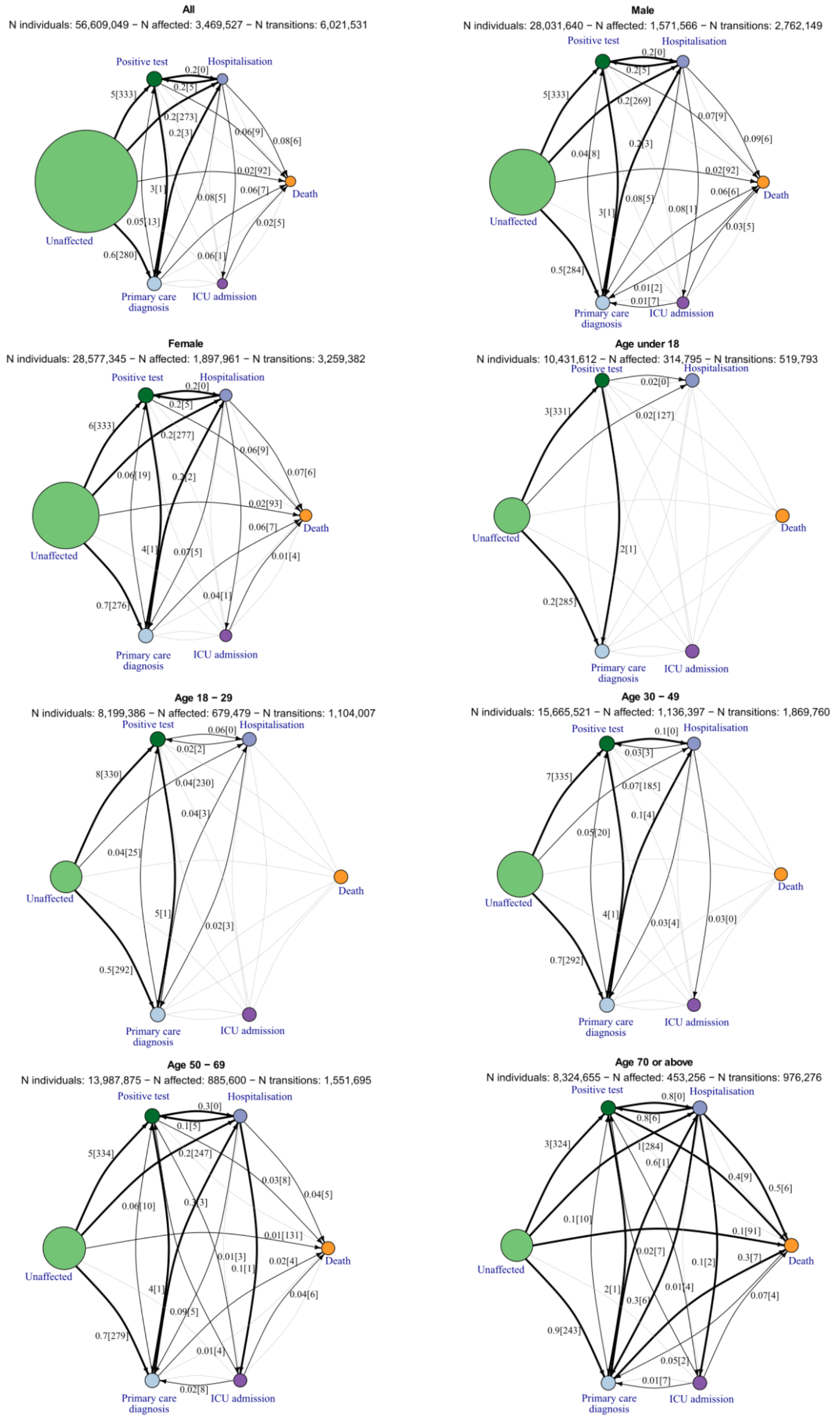


**Supplement figure 5: COVID-19 trajectory networks by gender and age groups.**
Networks show percentage of individuals transitioning and the median number of days passing between severity phenotypes stratified on sex and age groups. The size of the circles represent the number of individuals with that event relative to the total study population size of 56.6 million. Numbers on arrows are the percentage of individuals with the given transition (relative to N individuals in the group) and in square brackets median days between events across all individuals with that transition. Median days between unaffected and other severity phenotypes are larger, as these represent days from study start and the particular event. Thick arrows represent transitions occurring in ≥ 0.1%. Thin balck arrows represent transitions occurring in ≥ 0.01%. Any transitions occurring in fewer than 0.01% are not shown. N affected = N individuals with COVID-19.

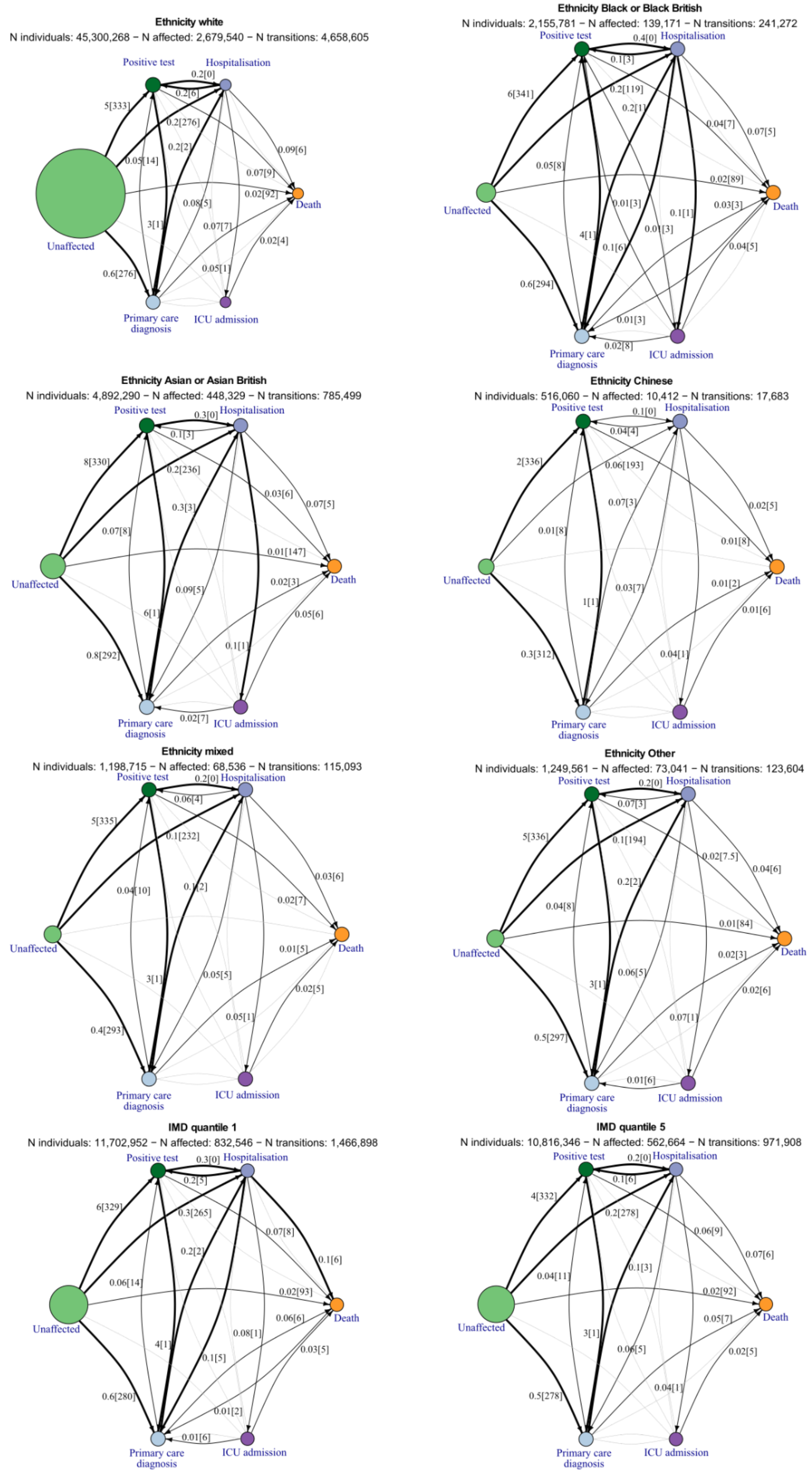


**Supplement figure 6: COVID-19 trajectory networks by ethnicity and IMD.**
Networks show showing percentage of individuals transitioning and the median number of days passing between severity phenotypes stratified on sex and age groups. The size of the circles represent the number of individuals with that event relative to the total study population size of 56.6 million. Numbers on arrows are the percentage of individuals with the given transition (relative to N individuals in the group) and in square brackets median days between events across all individuals with that transition. Median days between unaffected and other severity phenotypes are larger, as these represent days from study start and the particular event. Thick arrows represent transitions occurring in ≥ 0.1%. Thin balck arrows represent transitions occurring in ≥ 0.01%. Any transitions occurring in fewer than 0.01% are not shown. N affected = N individuals with COVID-19.

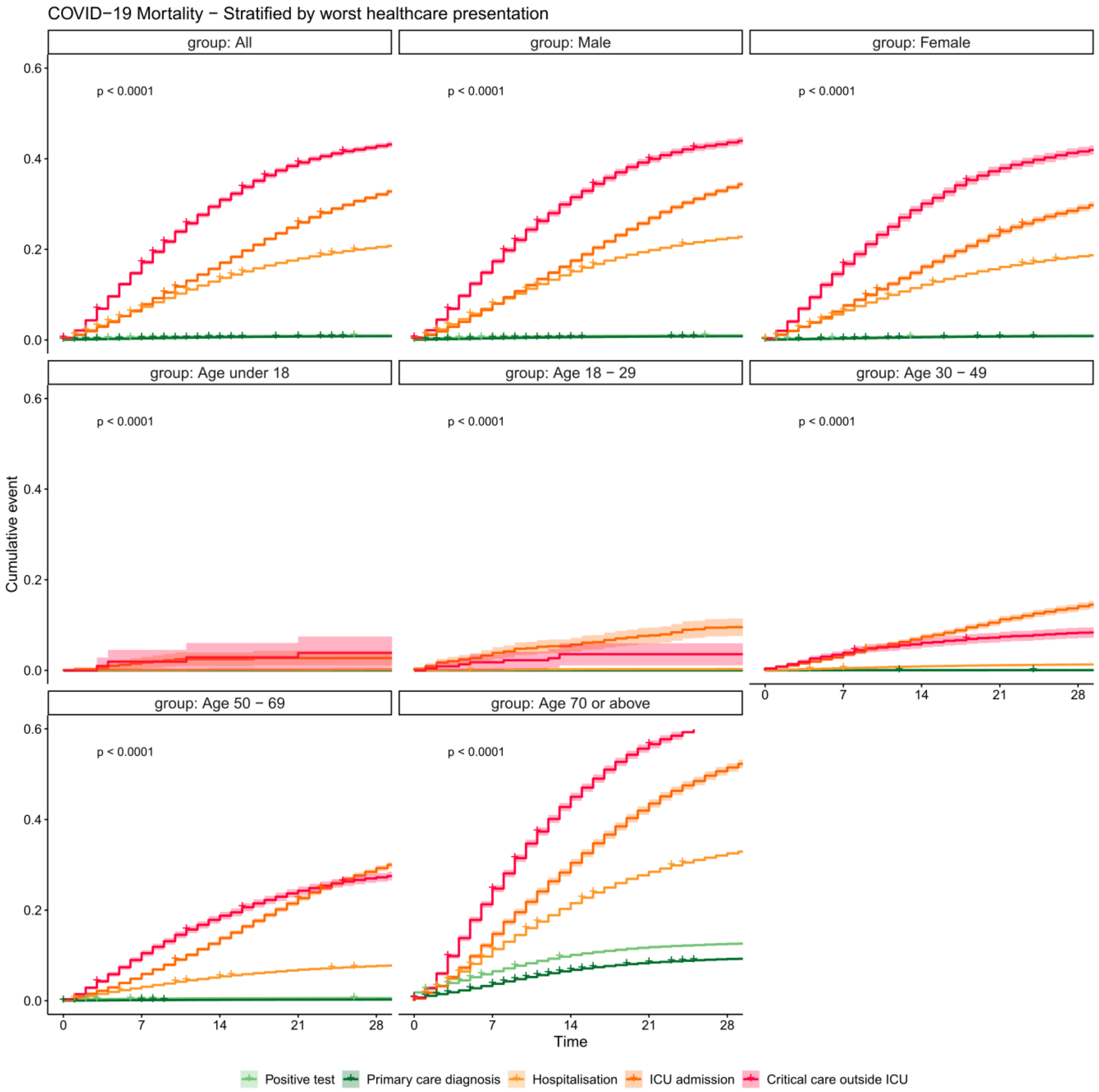


**Supplement figure 7: Kaplan Meier plot of COVID-19 mortality by gender and age groups.**
Curves are stratified by worst healthcare presentation, here listed in increasing order of severity; positive test (light green), primary care diagnosis (dark green), hospitalisation (orange) and ICU admission (blue). Note the ICU admission group does not include patients who received critical care outside of ICU wards. Shaded areas represent 95% confidence intervals in all panels.

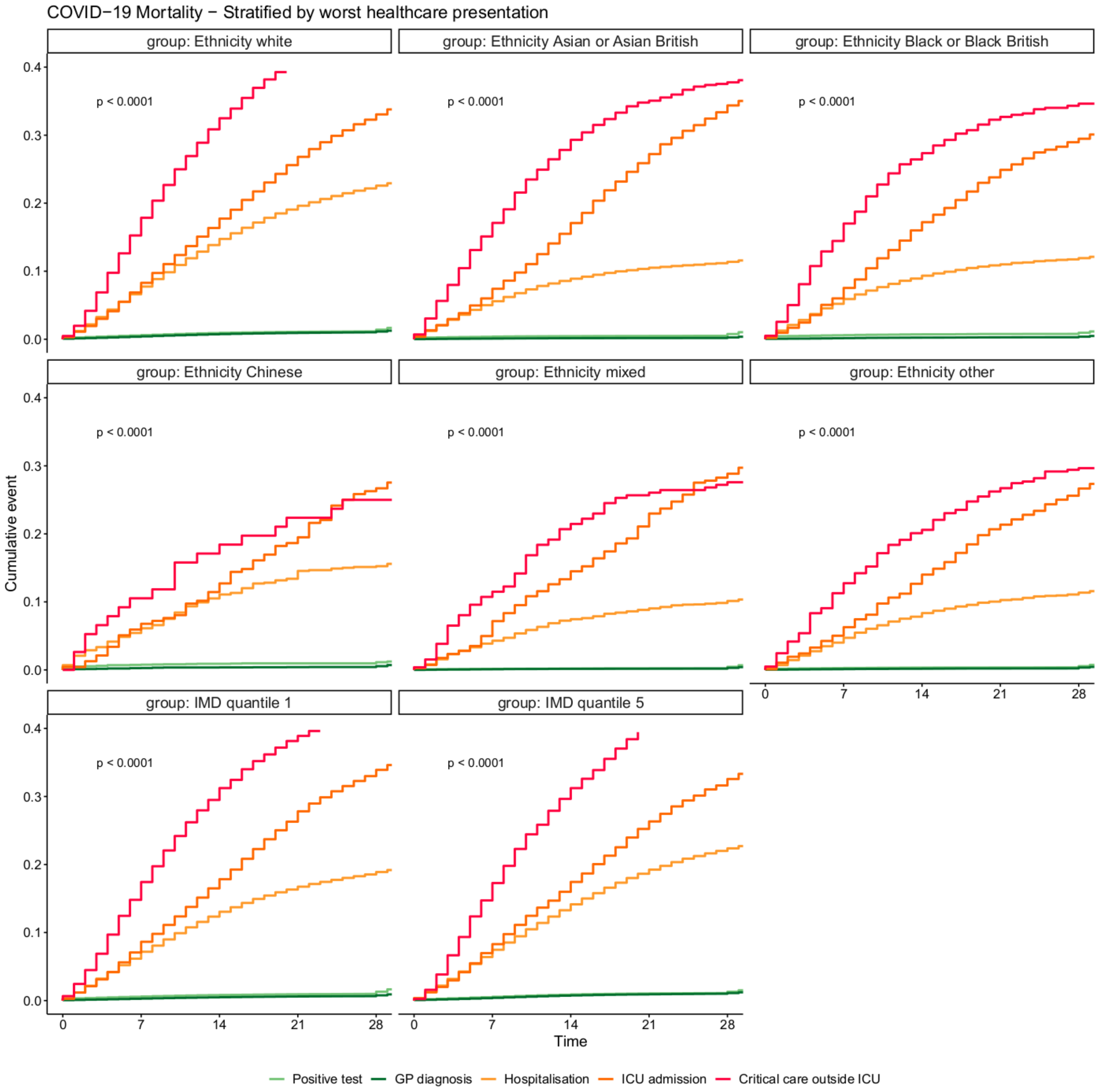


**Supplement figure 8: Kaplan Meier plot of COVID-19 mortality by ethnicity and IMD level.**
Curves are stratified by worst healthcare presentation, here listed in increasing order of severity; positive test (light green), primary care diagnosis (dark green), hospitalisation (orange) and ICU admission (blue). Note the ICU admission group does not include patients who received critical care outside of ICU wards. Shaded areas represent 95% confidence intervals in all panels.

#### Supplementary Tables

**Supplement table 1: Number of individuals identified from each data source stratified by COVID-19 event and as total individuals across all data sources.**

Date ranges shown for all included data sources after filtering as per cohort definition (see Figure 1).

| Data | SGSS | GDPPR | HES APC | SUS | CHESS | HES CC | Deaths | Total |
|---|---|---|---|---|---|---|---|---|
| Min. Date | 2020-01-24 | 2020-01-23 | 2020-01-23 | 2020-01-23 | 2020-01-23 | 2020-01-28 | 2020-01-30 | 2020-01-23 |
| Max. Date | 2021-05-31 | 2021-05-31 | 2021-05-31 | 2021-05-31 | 2021-05-31 | 2021-05-28 | 2021-05-31 | 2021-05-31 |
| Positive test | 3,114,784 | | | | | | | 3,114,784 |
| GP diagnosis | | 2,363,507 | | | | | | 2,363,507 |
| Hospitalisation | | | 353,061 | 359,751 | 52,082 | | | 364,260 |
| ECMO treatment | | | 416 | 520 | 312 | | | 549 |
| ICU admission | | | | | 15,151 | 35,846 | | 38,072 |
| IMV treatment | | | 16,212 | 16,500 | 7,613 | 18,415 | | 21,404 |
| NIV treatment | | | 42,108 | 42,992 | 12,011 | 26,959 | | 54,026 |
| Inpatient Death | | | 85,350 | 88,170 | | | | 88,675 |
| Fatal with diagnosis | | | | | | | 125,327 | 125,327 |
| Death without diagnosis | | | | | | | 13,083 | 13,083 |

**Supplement table 2: COVID-19 codes & frequencies.**

Codes appearing with a frequency <5 are masked as '<5' to protect privacy.

| COVID-19 Event | Code | Terminology | Description | Source | n |
|---|---|---|---|---|---|
| 01_Covid_positive_test | | | pillar_2 | SGSS | 2694960 |
| 01_Covid_positive_test | | | pillar_1 | SGSS | 741785 |
| 01_GP_covid_diagnosis | 1240581000000104 | SNOMED | Severe acute respiratory syndrome coronavirus 2 ribonucleic acid detected (finding) | GDPPR | 2127470 |
| 01_GP_covid_diagnosis | 1300721000000109 | SNOMED | Coronavirus disease 19 caused by severe acute respiratory syndrome coronavirus 2 confirmed by laboratory test (situation) | GDPPR | 419647 |
| 01_GP_covid_diagnosis | 1008541000000105 | SNOMED | Coronavirus ribonucleic acid detection assay (observable entity) | GDPPR | 257399 |
| 01_GP_covid_diagnosis | 1240751000000100 | SNOMED | Coronavirus disease 19 caused by severe acute respiratory syndrome coronavirus 2 (disorder) | GDPPR | 193545 |
| 01_GP_covid_diagnosis | 1321541000000108 | SNOMED | Severe acute respiratory syndrome coronavirus 2 immunoglobulin G detected (finding) | GDPPR | 67791 |
| 01_GP_covid_diagnosis | 1322781000000102 | SNOMED | Severe acute respiratory syndrome coronavirus 2 antigen detection result positive (finding) | GDPPR | 65011 |
| 01_GP_covid_diagnosis | 186747009 | SNOMED | Coronavirus infection (disorder) | GDPPR | 43078 |
| 01_GP_covid_diagnosis | 1300681000000102 | SNOMED | Assessment using coronavirus disease 19 severity scale (procedure) | GDPPR | 28030 |
| 01_GP_covid_diagnosis | 1300731000000106 | SNOMED | Coronavirus disease 19 caused by severe acute respiratory syndrome coronavirus 2 confirmed using clinical diagnostic criteria (situation) | GDPPR | 22156 |
| 01_GP_covid_diagnosis | 1240511000000106 | SNOMED | Detection of severe acute respiratory syndrome coronavirus 2 using polymerase chain reaction technique (procedure) | GDPPR | 16496 |
| 01_GP_covid_diagnosis | 1240551000000105 | SNOMED | Pneumonia caused by severe acute respiratory syndrome coronavirus 2 (disorder) | GDPPR | 12199 |
| 01_GP_covid_diagnosis | 1240741000000103 | SNOMED | Severe acute respiratory syndrome coronavirus 2 serology (observable entity) | GDPPR | 9873 |
| 01_GP_covid_diagnosis | 1321761000000103 | SNOMED | Severe acute respiratory syndrome coronavirus 2 immunoglobulin A detected (finding) | GDPPR | 7095 |
| 01_GP_covid_diagnosis | 1322871000000109 | SNOMED | Severe acute respiratory syndrome coronavirus 2 antibody detection result positive (finding) | GDPPR | 5568 |
| 01_GP_covid_diagnosis | 1300631000000101 | SNOMED | Coronavirus disease 19 severity score (observable entity) | GDPPR | 5054 |
| 01_GP_covid_diagnosis | 1240541000000107 | SNOMED | Infection of upper respiratory tract caused by severe acute respiratory syndrome coronavirus 2 (disorder) | GDPPR | 3421 |
| 01_GP_covid_diagnosis | 1029481000000103 | SNOMED | Coronavirus nucleic acid detection assay (observable entity) | GDPPR | 2578 |
| 01_GP_covid_diagnosis | 1300671000000104 | SNOMED | Coronavirus disease 19 severity scale (assessment scale) | GDPPR | 2059 |
| 01_GP_covid_diagnosis | 1321551000000106 | SNOMED | Severe acute respiratory syndrome coronavirus 2 immunoglobulin M detected (finding) | GDPPR | 1799 |
| 01_GP_covid_diagnosis | 1240401000000105 | SNOMED | Antibody to severe acute respiratory syndrome coronavirus 2 (substance) | GDPPR | 576 |

| | | | | | |
|---|---|---|---|---|---|
| 01_GP_covid_diagnosis | 1240421000000101 | SNOMED | Serotype severe acute respiratory syndrome coronavirus 2 (qualifier value) | GDPPR | 263 |
| 01_GP_covid_diagnosis | 1240571000000101 | SNOMED | Gastroenteritis caused by severe acute respiratory syndrome coronavirus 2 (disorder) | GDPPR | 230 |
| 01_GP_covid_diagnosis | 1240531000000103 | SNOMED | Myocarditis caused by severe acute respiratory syndrome coronavirus 2 (disorder) | GDPPR | 165 |
| 01_GP_covid_diagnosis | 1321341000000103 | SNOMED | Arbitrary concentration of severe acute respiratory syndrome coronavirus 2 immunoglobulin G in serum (observable entity) | GDPPR | 162 |
| 01_GP_covid_diagnosis | 1240391000000107 | SNOMED | Antigen of severe acute respiratory syndrome coronavirus 2 (substance) | GDPPR | 150 |
| 01_GP_covid_diagnosis | 1321331000000107 | SNOMED | Arbitrary concentration of severe acute respiratory syndrome coronavirus 2 total immunoglobulin in serum (observable entity) | GDPPR | 105 |
| 01_GP_covid_diagnosis | 1321301000000101 | SNOMED | Severe acute respiratory syndrome coronavirus 2 ribonucleic acid qualitative existence in specimen (observable entity) | GDPPR | 99 |
| 01_GP_covid_diagnosis | 1240561000000108 | SNOMED | Encephalopathy caused by severe acute respiratory syndrome coronavirus 2 (disorder) | GDPPR | 70 |
| 01_GP_covid_diagnosis | 1321241000000105 | SNOMED | Cardiomyopathy caused by severe acute respiratory syndrome coronavirus 2 (disorder) | GDPPR | 23 |
| 01_GP_covid_diagnosis | 1321351000000100 | SNOMED | Arbitrary concentration of severe acute respiratory syndrome coronavirus 2 immunoglobulin M in serum (observable entity) | GDPPR | 21 |
| 01_GP_covid_diagnosis | 1240521000000100 | SNOMED | Otitis media caused by severe acute respiratory syndrome coronavirus 2 (disorder) | GDPPR | 17 |
| 01_GP_covid_diagnosis | 1321321000000105 | SNOMED | Severe acute respiratory syndrome coronavirus 2 immunoglobulin G qualitative existence in specimen (observable entity) | GDPPR | 15 |
| 01_GP_covid_diagnosis | 1321311000000104 | SNOMED | Severe acute respiratory syndrome coronavirus 2 immunoglobulin M qualitative existence in specimen (observable entity) | GDPPR | 11 |
| 01_GP_covid_diagnosis | 1321811000000105 | SNOMED | Severe acute respiratory syndrome coronavirus 2 immunoglobulin A qualitative existence in specimen (observable entity) | GDPPR | <5 |
| 01_GP_covid_diagnosis | 120814005 | SNOMED | Coronavirus antibody (substance) | GDPPR | <5 |
| 01_GP_covid_diagnosis | 1321201000000107 | SNOMED | Coronavirus disease 19 caused by severe acute respiratory syndrome coronavirus 2 health issues simple reference set (foundation metadata concept) | GDPPR | <5 |
| 01_GP_covid_diagnosis | 1321181000000108 | SNOMED | Coronavirus disease 19 caused by severe acute respiratory syndrome coronavirus 2 record extraction simple reference set (foundation metadata concept) | GDPPR | <5 |
| 01_GP_covid_diagnosis | 1240381000000105 | SNOMED | Severe acute respiratory syndrome coronavirus 2 (organism) | GDPPR | <5 |
| 01_GP_covid_diagnosis | 1321801000000108 | SNOMED | Arbitrary concentration of severe acute respiratory syndrome coronavirus 2 immunoglobulin A in serum (observable entity) | GDPPR | <5 |
| 01_GP_covid_diagnosis | 1240411000000107 | SNOMED | Ribonucleic acid of severe acute respiratory syndrome coronavirus 2 (substance) | GDPPR | <5 |
| 01_GP_covid_diagnosis | 1321191000000105 | SNOMED | Coronavirus disease 19 caused by severe acute respiratory syndrome coronavirus 2 procedures simple reference set (foundation metadata concept) | GDPPR | <5 |
| 02_Covid_admission | U07.1 | ICD10 | Confirmed_COVID19 | SUS | 667110 |
| 02_Covid_admission | U07.1 | ICD10 | Confirmed_COVID19 | HES APC | 654497 |
| 02_Covid_admission | U07.2 | ICD10 | Suspected_COVID19 | SUS | 64012 |
| 02_Covid_admission | U07.2 | ICD10 | Suspected_COVID19 | HES APC | 58567 |
| 02_Covid_admission | | | HospitalAdmissionDate IS NOT null | CHESS | 55454 |
| 03_ECMO_treatment | X58.1 | OPCS | Extracorporeal membrane oxygenation | SUS | 588 |
| 03_ECMO_treatment | X58.1 | OPCS | Extracorporeal membrane oxygenation | HES APC | 461 |
| 03_ECMO_treatment | | | RespiratorySupportECMO == Yes | CHESS | 379 |
| 03_ICU_admission | | | id is in hes_cc table | HES CC | 44990 |
| 03_ICU_admission | | | DateAdmittedICU IS NOT null | CHESS | 16291 |
| 03_IMV_treatment | | | ARESSUPDAYS > 0 | HES CC | 22835 |
| 03_IMV_treatment | E85.1 | OPCS | Invasive ventilation | SUS | 18716 |
| 03_IMV_treatment | E85.1 | OPCS | Invasive ventilation | HES APC | 18018 |
| 03_IMV_treatment | | | Invasivemechanicalventilation == Yes | CHESS | 8382 |
| 03_IMV_treatment | X56 | OPCS | Intubation of trachea | SUS | 451 |
| 03_IMV_treatment | X56 | OPCS | Intubation of trachea | HES APC | 414 |
| 03_NIV_treatment | E85.6 | OPCS | Continuous positive airway pressure | SUS | 36857 |
| 03_NIV_treatment | E85.6 | OPCS | Continuous positive airway pressure | HES APC | 36175 |
| 03_NIV_treatment | | | bressupdays > 0 | HES CC | 30785 |
| 03_NIV_treatment | E85.2 | OPCS | Non-invasive ventilation NEC | SUS | 17622 |
| 03_NIV_treatment | E85.2 | OPCS | Non-invasive ventilation NEC | HES APC | 16242 |
| 03_NIV_treatment | | | Highflownasaloxygen OR NoninvasiveMechanicalventilation == Yes | CHESS | 12280 |
| 04_Covid_inpatient_death | | | DISCHARGE_METHOD_HOSPITAL_PROVIDER_SPELL = 4 (Died) | SUS | 89811 |
| 04_Covid_inpatient_death | | | DISMETH = 4 (Died) | HES APC | 85328 |
| 04_Covid_inpatient_death | | | DISCHARGE_DESTINATION_HOSPITAL_PROVIDER_SPELL = 79 (Not applicable - PATIENT died or still birth) | SUS | 155 |
| 04_Covid_inpatient_death | | | DISDEST = 79 (Not applicable - PATIENT died or still birth) | HES APC | 108 |

| | | | | | |
|---|---|---|---|---|---|
| 04_Fatal_with_covid_diagnosis | U071 | ICD10 | | deaths | 123154 |
| 04_Fatal_with_covid_diagnosis | U072 | ICD10 | | deaths | 4032 |
| 04_Fatal_without_covid_diagnosis | | | ONS death within 28 days | deaths | 13085 |

**Supplement table 3: 270 CALIBER phenotypes, aggregated into 16 categories**
Table shows the number of individuals within the study cohort identified from GDPPR (SNOMED-CT) and HES APC (ICD-10, OPCS-4).

| Category | Phenotype | Individuals |
|---|---|---:|
| benign_neoplasm_cin | benign_neoplasm_of_colon_rectum_anus_and_anal_... | 82,933 |
| benign_neoplasm_cin | benign_neoplasm_of_ovary | 53,602 |
| benign_neoplasm_cin | benign_neoplasm_and_polyp_of_uterus | 30,807 |
| benign_neoplasm_cin | benign_neoplasm_of_stomach_and_duodenum | 25,003 |
| benign_neoplasm_cin | haemangioma_any_site | 10,885 |
| benign_neoplasm_cin | carcinoma_in_situ_cervical | 4,351 |
| benign_neoplasm_cin | benign_neoplasm_of_brain_and_other_parts_of_ce... | 2,665 |
| cancers | myelodysplastic_syndromes | 284,874 |
| cancers | primary_malignancy_other_organs | 231,644 |
| cancers | primary_malignancy_other_skin_and_subcutaneous... | 220,826 |
| cancers | primary_malignancy_breast | 104,193 |
| cancers | primary_malignancy_cervical | 46,985 |
| cancers | primary_malignancy_prostate | 19,692 |
| cancers | primary_malignancy_colorectal_and_anus | 14,410 |
| cancers | primary_malignancy_malignant_melanoma | 9,706 |
| cancers | non-hodgkin_lymphoma | 7,830 |
| cancers | secondary_malignancy_other_organs | 7,438 |
| cancers | primary_malignancy_bladder | 7,001 |
| cancers | monoclonal_gammopathy_of_undetermined_signific... | 6,665 |
| cancers | leukaemia | 6,051 |
| cancers | hodgkin_lymphoma | 5,570 |
| cancers | multiple_myeloma_and_malignant_plasma_cell_neo... | 5,301 |
| cancers | primary_malignancy_lung_and_trachea | 4,401 |
| cancers | secondary_malignancy_bone | 4,272 |
| cancers | primary_malignancy_kidney_and_ureter | 3,438 |
| cancers | primary_malignancy_uterine | 3,011 |
| cancers | secondary_malignancy_lung | 2,875 |
| cancers | secondary_malignancy_liver_and_intrahepatic_bi... | 2,643 |
| cancers | primary_malignancy_testicular | 2,442 |
| cancers | primary_malignancy_ovarian | 2,300 |
| cancers | primary_malignancy_oro-pharyngeal | 2,097 |
| cancers | polycythaemia_vera | 2,049 |
| cancers | primary_malignancy_thyroid | 1,871 |
| cancers | secondary_malignancy_retroperitoneum_and_perit... | 1,536 |
| cancers | primary_malignancy_oesophageal | 1,039 |
| cancers | primary_malignancy_brain_other_cns_and_intracr... | 1,006 |
| cancers | secondary_malignancy_lymph_nodes | 780 |
| cancers | primary_malignancy_liver | 748 |
| cancers | secondary_malignancy_brain_other_cns_and_intra... | 746 |
| cancers | primary_malignancy_stomach | 720 |
| cancers | secondary_malignancy_pleura | 564 |
| cancers | primary_malignancy_pancreatic | 506 |
| cancers | primary_malignancy_bone_and_articular_cartilage | 399 |
| cancers | secondary_malignancy_bowel | 397 |
| cancers | primary_malignancy_biliary_tract | 313 |
| cancers | secondary_malignancy_adrenal_gland | 303 |
| cancers | primary_malignancy_mesothelioma | 68 |
| cancers | primary_malignancy_multiple_independent_sites | 8 |
| diseases_of_the_circulatory_system | hypertension | 568,174 |
| diseases_of_the_circulatory_system | coronary_heart_disease_not_otherwise_specified | 161,781 |
| diseases_of_the_circulatory_system | stable_angina | 110,546 |
| diseases_of_the_circulatory_system | atrial_fibrillation | 102,862 |
| diseases_of_the_circulatory_system | myocardial_infarction | 88,821 |
| diseases_of_the_circulatory_system | heart_failure | 58,762 |
| diseases_of_the_circulatory_system | stroke_nos | 51,193 |

| | | |
|---|---|---|
| diseases_of_the_circulatory_system | transient_ischaemic_attack | 47,975 |
| diseases_of_the_circulatory_system | ischaemic_stroke | 45,939 |
| diseases_of_the_circulatory_system | peripheral_arterial_disease | 43,035 |
| diseases_of_the_circulatory_system | unstable_angina | 40,536 |
| diseases_of_the_circulatory_system | supraventricular_tachycardia | 26,549 |
| diseases_of_the_circulatory_system | venous_thromboembolic_disease_excl_pe | 23,853 |
| diseases_of_the_circulatory_system | right_bundle_branch_block | 23,775 |
| diseases_of_the_circulatory_system | left_bundle_branch_block | 20,374 |
| diseases_of_the_circulatory_system | atrioventricular_block_first_degree | 18,001 |
| diseases_of_the_circulatory_system | abdominal_aortic_aneurysm | 12,956 |
| diseases_of_the_circulatory_system | raynauds_syndrome | 8,368 |
| diseases_of_the_circulatory_system | secondary_pulmonary_hypertension | 7,782 |
| diseases_of_the_circulatory_system | other_cardiomyopathy | 7,318 |
| diseases_of_the_circulatory_system | atrioventricular_block_complete | 7,014 |
| diseases_of_the_circulatory_system | pericardial_effusion_noninflammatory | 6,956 |
| diseases_of_the_circulatory_system | intracerebral_haemorrhage | 6,031 |
| diseases_of_the_circulatory_system | ventricular_tachycardia | 5,824 |
| diseases_of_the_circulatory_system | atrioventricular_block_second_degree | 5,009 |
| diseases_of_the_circulatory_system | dilated_cardiomyopathy | 4,659 |
| diseases_of_the_circulatory_system | sick_sinus_syndrome | 4,226 |
| diseases_of_the_circulatory_system | subdural_haematoma_-_nontraumatic | 3,302 |
| diseases_of_the_circulatory_system | primary_pulmonary_hypertension | 3,139 |
| diseases_of_the_circulatory_system | trifascicular_block | 2,015 |
| diseases_of_the_circulatory_system | bifascicular_block | 1,992 |
| diseases_of_the_circulatory_system | hypertrophic_cardiomyopathy | 1,897 |
| diseases_of_the_circulatory_system | subarachnoid_haemorrhage | 863 |
| diseases_of_the_circulatory_system | pulmonary_embolism | 433 |
| diseases_of_the_circulatory_system | rheumatic_valve_dz | 140 |
| diseases_of_the_digestive_system | abdominal_hernia | 115,615 |
| diseases_of_the_digestive_system | oesophagitis_and_oesophageal_ulcer | 72,309 |
| diseases_of_the_digestive_system | appendicitis | 62,962 |
| diseases_of_the_digestive_system | cholecystitis | 52,391 |
| diseases_of_the_digestive_system | fatty_liver | 28,930 |
| diseases_of_the_digestive_system | anal_fissure | 18,229 |
| diseases_of_the_digestive_system | barretts_oesophagus | 17,253 |
| diseases_of_the_digestive_system | peritonitis | 16,205 |
| diseases_of_the_digestive_system | coeliac_disease | 15,883 |
| diseases_of_the_digestive_system | liver_fibrosis_sclerosis_and_cirrhosis | 12,408 |
| diseases_of_the_digestive_system | anorectal_fistula | 12,332 |
| diseases_of_the_digestive_system | cholangitis | 7,607 |
| diseases_of_the_digestive_system | pancreatitis | 7,509 |
| diseases_of_the_digestive_system | anorectal_prolapse | 7,076 |
| diseases_of_the_digestive_system | portal_hypertension | 5,661 |
| diseases_of_the_digestive_system | gastro-oesophageal_reflux_disease | 4,892 |
| diseases_of_the_digestive_system | volvulus | 4,277 |
| diseases_of_the_digestive_system | angiodysplasia_of_colon | 3,155 |
| diseases_of_the_digestive_system | autoimmune_liver_disease | 2,894 |
| diseases_of_the_digestive_system | oesophageal_varices | 2,709 |
| diseases_of_the_digestive_system | alcoholic_liver_disease | 2,163 |
| diseases_of_the_digestive_system | diaphragmatic_hernia | 2,067 |
| diseases_of_the_digestive_system | hepatic_failure | 1,464 |
| diseases_of_the_digestive_system | peptic_ulcer_disease | 1,381 |
| diseases_of_the_digestive_system | diverticular_disease_of_intestine_acute_and_ch... | 184 |
| diseases_of_the_ear | hearing_loss | 5,970 |
| diseases_of_the_ear | tinnitus | 5,063 |
| diseases_of_the_ear | meniere_disease | 3,930 |
| diseases_of_the_endocrine_system | diabetes | 280,896 |
| diseases_of_the_endocrine_system | obesity | 203,250 |
| diseases_of_the_endocrine_system | hypo_or_hyperthyroidism | 171,787 |
| diseases_of_the_endocrine_system | polycystic_ovarian_syndrome | 24,459 |
| diseases_of_the_endocrine_system | syndrome_of_inappropriate_secretion_of_antidiu... | 19,025 |
| diseases_of_the_endocrine_system | hyperparathyroidism | 7,433 |
| diseases_of_the_endocrine_system | cystic_fibrosis | 864 |
| diseases_of_the_eye | diabetic_ophthalmic_complications | 202,348 |
| diseases_of_the_eye | cataract | 177,536 |
| diseases_of_the_eye | glaucoma | 33,775 |
| diseases_of_the_eye | macular_degeneration | 32,503 |

| | | |
|---|---|---|
| diseases_of_the_eye | retinal_detachments_and_breaks | 15,994 |
| diseases_of_the_eye | ptosis_of_eyelid | 7,665 |
| diseases_of_the_eye | anterior_and_intermediate_uveitis | 3,172 |
| diseases_of_the_eye | keratitis | 567 |
| diseases_of_the_eye | scleritis_and_episcleritis | 449 |
| diseases_of_the_eye | posterior_uveitis | 260 |
| diseases_of_the_eye | retinal_vascular_occlusions | 99 |
| diseases_of_the_eye | visual_impairment_and_blindness | 54 |
| diseases_of_the_genitourinary_system | acute_kidney_injury | 106,099 |
| diseases_of_the_genitourinary_system | menorrhagia_and_polymenorrhoea | 85,377 |
| diseases_of_the_genitourinary_system | urolithiasis | 58,758 |
| diseases_of_the_genitourinary_system | obstructive_and_reflux_uropathy | 29,746 |
| diseases_of_the_genitourinary_system | urinary_incontinence | 29,696 |
| diseases_of_the_genitourinary_system | postmenopausal_bleeding | 23,098 |
| diseases_of_the_genitourinary_system | end_stage_renal_disease | 17,055 |
| diseases_of_the_genitourinary_system | dysmenorrhoea | 16,493 |
| diseases_of_the_genitourinary_system | glomerulonephritis | 15,858 |
| diseases_of_the_genitourinary_system | hydrocoele_incl_infected | 14,197 |
| diseases_of_the_genitourinary_system | endometrial_hyperplasia_and_hypertrophy | 13,294 |
| diseases_of_the_genitourinary_system | postcoital_and_contact_bleeding | 12,034 |
| diseases_of_the_genitourinary_system | non-acute_cystitis | 5,603 |
| diseases_of_the_genitourinary_system | erectile_dysfunction | 2,680 |
| diseases_of_the_genitourinary_system | tubulo-interstitial_nephritis | 1,408 |
| diseases_of_the_genitourinary_system | undescended_testicle | 33 |
| diseases_of_the_genitourinary_system | male_infertility | 13 |
| diseases_of_the_respiratory_system | asthma | 528,582 |
| diseases_of_the_respiratory_system | copd | 85,731 |
| diseases_of_the_respiratory_system | allergic_and_chronic_rhinitis | 68,720 |
| diseases_of_the_respiratory_system | chronic_sinusitis | 60,075 |
| diseases_of_the_respiratory_system | sleep_apnoea | 41,581 |
| diseases_of_the_respiratory_system | pulmonary_collapse_excl_pneumothorax | 30,531 |
| diseases_of_the_respiratory_system | hypertrophy_of_nasal_turbinates | 18,848 |
| diseases_of_the_respiratory_system | bronchiectasis | 17,719 |
| diseases_of_the_respiratory_system | aspiration_pneumonitis | 13,829 |
| diseases_of_the_respiratory_system | other_interstitial_pulmonary_diseases_with_fib... | 10,684 |
| diseases_of_the_respiratory_system | pleural_plaque | 6,469 |
| diseases_of_the_respiratory_system | respiratory_failure | 2,013 |
| diseases_of_the_respiratory_system | asbestosis | 803 |
| diseases_of_the_respiratory_system | pleural_effusion | 94 |
| diseases_of_the_respiratory_system | pneumothorax | 25 |
| diseases_of_the_respiratory_system | nasal_polyp | 0 |
| haematological_immunological_conditions | vitamin_b12_deficiency_anaemia | <5 |
| haematological_immunological_conditions | secondary_or_other_thrombocytopaenia | 14,326 |
| haematological_immunological_conditions | splenomegaly | 9,774 |
| haematological_immunological_conditions | sickle-cell_trait | 6,587 |
| haematological_immunological_conditions | thalassaemia_trait | 5,871 |
| haematological_immunological_conditions | hyposplenism | 4,498 |
| haematological_immunological_conditions | primary_or_idiopathic_thrombocytopaenia | 4,208 |
| haematological_immunological_conditions | thalassaemia | 3,606 |
| haematological_immunological_conditions | thrombophilia | 3,570 |
| haematological_immunological_conditions | agranulocytosis | 3,304 |
| haematological_immunological_conditions | sickle-cell_anaemia | 2,816 |
| haematological_immunological_conditions | secondary_polycythaemia | 2,635 |
| haematological_immunological_conditions | aplastic_anaemias | 2,196 |
| haematological_immunological_conditions | immunodeficiencies | 1,984 |
| haematological_immunological_conditions | sarcoidosis | 1,520 |
| haematological_immunological_conditions | other_haemolytic_anaemias | 383 |
| haematological_immunological_conditions | other_anaemias | 179 |
| haematological_immunological_conditions | iron_deficiency_anaemia | 65 |
| infectious_diseases | bacterial_diseases_excl_tb | 238,377 |
| infectious_diseases | other_or_unspecified_infectious_organisms | 238,106 |
| infectious_diseases | hiv | 219,038 |
| infectious_diseases | urinary_tract_infections | 199,628 |
| infectious_diseases | viral_diseases_excl_chronic_hepatitis_hiv | 65,332 |
| infectious_diseases | infections_of_other_or_unspecified_organs | 56,208 |
| infectious_diseases | lower_respiratory_tract_infections | 53,756 |
| infectious_diseases | infection_of_other_or_unspecified_genitourinar... | 20,132 |

| | | |
|---|---|---|
| infectious_diseases | infection_of_skin_and_subcutaneous_tissues | 17,584 |
| infectious_diseases | infections_of_the_digestive_system | 17,376 |
| infectious_diseases | other_nervous_system_infections | 9,924 |
| infectious_diseases | infection_of_male_genital_system | 6,650 |
| infectious_diseases | eye_infections | 6,574 |
| infectious_diseases | infections_of_the_heart | 3,877 |
| infectious_diseases | infection_of_bones_and_joints | 3,861 |
| infectious_diseases | infection_of_anal_and_rectal_regions | 3,435 |
| infectious_diseases | infection_of_liver | 3,045 |
| infectious_diseases | chronic_viral_hepatitis | 2,543 |
| infectious_diseases | parasitic_infections | 2,027 |
| infectious_diseases | mycoses | 1,856 |
| infectious_diseases | ear_and_upper_respiratory_tract_infections | 1,783 |
| infectious_diseases | meningitis | 1,763 |
| infectious_diseases | septicaemia | 1,572 |
| infectious_diseases | tuberculosis | 1,216 |
| infectious_diseases | encephalitis | 538 |
| infectious_diseases | rheumatic_fever | 239 |
| mental_health_disorders | depression | 638,222 |
| mental_health_disorders | bipolar_affective_disorder_and_mania | 396,659 |
| mental_health_disorders | anxiety_disorders | 261,938 |
| mental_health_disorders | other_psychoactive_substance_misuse | 135,585 |
| mental_health_disorders | alcohol_problems | 96,206 |
| mental_health_disorders | dementia | 91,367 |
| mental_health_disorders | intellectual_disability | 39,980 |
| mental_health_disorders | schizophrenia_schizotypal_and_delusional_disor... | 23,940 |
| mental_health_disorders | autism_and_aspergers_syndrome | 23,504 |
| mental_health_disorders | hyperkinetic_disorders | 14,624 |
| mental_health_disorders | anorexia_and_bulimia_nervosa | 3,046 |
| mental_health_disorders | personality_disorders | 201 |
| mental_health_disorders | delirium_not_induced_by_alcohol_and_other_psyc... | 82 |
| musculoskeletal_conditions | osteoporosis | 73,899 |
| musculoskeletal_conditions | spondylosis | 66,395 |
| musculoskeletal_conditions | carpal_tunnel_syndrome | 51,553 |
| musculoskeletal_conditions | fracture_of_wrist | 44,024 |
| musculoskeletal_conditions | enthesopathies__synovial_disorders | 42,187 |
| musculoskeletal_conditions | fracture_of_hip | 37,679 |
| musculoskeletal_conditions | spinal_stenosis | 29,367 |
| musculoskeletal_conditions | rheumatoid_arthritis | 26,342 |
| musculoskeletal_conditions | intervertebral_disc_disorders | 17,427 |
| musculoskeletal_conditions | polymyalgia_rheumatica | 12,729 |
| musculoskeletal_conditions | collapsed_vertebra | 9,801 |
| musculoskeletal_conditions | spondylolisthesis | 9,590 |
| musculoskeletal_conditions | fibromatoses | 9,186 |
| musculoskeletal_conditions | psoriatic_arthropathy | 4,769 |
| musculoskeletal_conditions | giant_cell_arteritis | 3,233 |
| musculoskeletal_conditions | sjogrens_disease | 2,627 |
| musculoskeletal_conditions | enteropathic_arthropathy | 462 |
| musculoskeletal_conditions | gout | 46 |
| musculoskeletal_conditions | systemic_sclerosis | 12 |
| musculoskeletal_conditions | lupus_erythematosus_local_and_systemic | 0 |
| neurological_conditions | epilepsy | 54,629 |
| neurological_conditions | postviral_fatigue_syndrome_neurasthenia_and_fi... | 31,424 |
| neurological_conditions | peripheral_neuropathies_excluding_cranial_nerv... | 28,445 |
| neurological_conditions | diabetic_neurological_complications | 23,834 |
| neurological_conditions | parkinsons_disease | 13,097 |
| neurological_conditions | bells_palsy | 8,268 |
| neurological_conditions | disorders_of_autonomic_nervous_system | 3,624 |
| neurological_conditions | intracranial_hypertension | 3,226 |
| neurological_conditions | trigeminal_neuralgia | 3,178 |
| neurological_conditions | essential_tremor | 2,934 |
| neurological_conditions | myasthenia_gravis | 1,533 |
| neurological_conditions | motor_neuron_disease | 951 |
| perinatal_conditions | slow_fetal_growth_or_low_birth_weight | 24,016 |
| perinatal_conditions | prematurity | 15,232 |
| perinatal_conditions | congenital_malformations_of_cardiac_septa | 13,995 |
| perinatal_conditions | high_birth_weight | 6,720 |

| | | |
|---|---|---|
| perinatal_conditions | post-term_infant | 3,426 |
| perinatal_conditions | patent_ductus_arteriosus | 2,857 |
| perinatal_conditions | downs_syndrome | 2,713 |
| perinatal_conditions | intrauterine_hypoxia | 1,143 |
| perinatal_conditions | spina_bifida | 896 |
| skin_conditions | seborrheic_dermatitis | <5 |
| skin_conditions | dermatitis_atopc_contact_other_unspecified | 70,563 |
| skin_conditions | pilonidal_cyst_sinus | 15,805 |
| skin_conditions | actinic_keratosis | 11,061 |
| skin_conditions | psoriasis | 4,603 |
| skin_conditions | hidradenitis_suppurativa | 3,261 |
| skin_conditions | acne | 2,036 |
| skin_conditions | rosacea | 1,435 |
| skin_conditions | lichen_planus | 67 |

**Supplement table 4: Demographic overview of deceased patients**

Table is stratified by individuals identified with or without a formal COVID-19 diagnosis listed on the death certificate, and/or COVID-19 inpatient deaths, as compared with the total population of all patients with a COVID-19 event.

| | **Fatal with COVID-19 diagnosis** | **Fatal without COVID-19 diagnosis** | **COVID-19 Inpatient death** | **COVID-19 Death no hospital contact** | **COVID-19 Death no hospital contact - Wave 1** | **COVID-19 Death no hospital contact - Wave 2** | **All COVID events** |
|---|---|---|---|---|---|---|---|
| n | 125327 (3.6) | 13083 (0.4) | 88675 (2.6) | 39510 (1.1) | 16325 (6.2) | 19951 (0.7) | 3469528 (100) |
| **Sex** | | | | | | | |
| Female | 57287 (45.7) | 6350 (48.5) | 36424 (41.1) | 21971 (55.6) | 8830 (54.1) | 11546 (57.9) | 1897963 (54.7) |
| Unknown | 0 (0) | 0 (0) | 0 (0) | 0 (0) | 0 (0) | 0 (0) | 0 (0) |
| **Age** | | | | | | | |
| Under 18 | 20 (0) | 20 (0.2) | 27 (0) | 19 (0) | 6 (0) | 10 (0.1) | 337664 (9.7) |
| Age 18 - 29 | 165 (0.1) | 38 (0.3) | 136 (0.2) | 70 (0.2) | 27 (0.2) | 35 (0.2) | 694821 (20) |
| Age 30 - 49 | 2328 (1.9) | 357 (2.7) | 1864 (2.1) | 733 (1.9) | 217 (1.3) | 416 (2.1) | 1134935 (32.7) |
| Age 50 - 69 | 18465 (14.7) | 2035 (15.6) | 15769 (17.8) | 3867 (9.8) | 1295 (7.9) | 2036 (10.2) | 862436 (24.9) |
| >= 70 | 104349 (83.3) | 10633 (81.3) | 70879 (79.9) | 34821 (88.1) | 14780 (90.5) | 17454 (87.5) | 439672 (12.7) |
| Unknown | 0 (0) | 0 (0) | 0 (0) | 0 (0) | 0 (0) | 0 (0) | 0 (0) |
| **Ethnicity** | | | | | | | |
| White | 109568 (87.4) | 11972 (91.5) | 76036 (85.7) | 36092 (91.3) | 14947 (91.6) | 18205 (91.2) | 2679545 (77.2) |
| Asian or asian british | 8569 (6.8) | 584 (4.5) | 7109 (8) | 1572 (4) | 513 (3.1) | 885 (4.4) | 448341 (12.9) |
| Black or black british | 3763 (3) | 248 (1.9) | 2972 (3.4) | 869 (2.2) | 412 (2.5) | 414 (2.1) | 139152 (4) |
| Chinese | 303 (0.2) | 23 (0.2) | 229 (0.3) | 85 (0.2) | 42 (0.3) | 38 (0.2) | 10417 (0.3) |
| Mixed and others | 2221 (1.8) | 179 (1.4) | 1755 (2) | 521 (1.3) | 257 (1.6) | 221 (1.1) | 141580 (4.1) |
| Unknown ethnicity | 903 (0.7) | 77 (0.6) | 574 (0.6) | 371 (0.9) | 154 (0.9) | 188 (0.9) | 50493 (1.5) |
| IMD Fifths (%) | | | | | | | |
| 1 (most deprived) | 29198 (23.3) | 2992 (22.9) | 21821 (24.6) | 7892 (20) | 3380 (20.7) | 3849 (19.3) | 832581 (24) |
| 5 (least deprived) | 21018 (16.8) | 2330 (17.8) | 14002 (15.8) | 7651 (19.4) | 3201 (19.6) | 3922 (19.7) | 562638 (16.2) |
| Unknown | 111 (0.1) | 12 (0.1) | 58 (0.1) | 53 (0.1) | 25 (0.2) | 23 (0.1) | 2911 (0.1) |
| **COVID-19 events** | | | | | | | |
| COVID-19 positive test | 106941 (85.3) | 5365 (41) | 79670 (89.8) | 23567 (59.6) | 6405 (39.2) | 15913 (79.8) | 3114784 (89.8) |
| GP COVID-19 diagnosis | 52286 (41.7) | 5029 (38.4) | 31525 (35.6) | 19527 (49.4) | 5933 (36.3) | 11563 (58) | 2363507 (68.1) |
| COVID-19 admission | 91308 (72.9) | 7593 (58) | 88657 (100) | 0 (0) | 0 (0) | 0 (0) | 364260 (10.5) |
| ICU admission | 14429 (11.5) | 619 (4.7) | 15074 (17) | 0 (0) | 0 (0) | 0 (0) | 38072 (1.1) |
| NIV treatment | 21899 (17.5) | 773 (5.9) | 22498 (25.4) | 0 (0) | 0 (0) | 0 (0) | 54026 (1.6) |
| IMV treatment | 10596 (8.5) | 422 (3.2) | 11153 (12.6) | 0 (0) | 0 (0) | 0 (0) | 21404 (0.6) |
| ECMO treatment | 194 (0.2) | 1 (0) | 204 (0.2) | 0 (0) | 0 (0) | 0 (0) | 549 (0) |
| Fatal with COVID-19 diagnosis | 125327 (100) | 0 (0) | 80786 (91.1) | 34018 (86.1) | 15100 (92.5) | 17310 (86.8) | 125327 (3.6) |
| Fatal without COVID-19 diagnosis | 0 (0) | 13083 (100) | 5391 (6.1) | 5490 (13.9) | 1223 (7.5) | 2641 (13.2) | 13083 (0.4) |
| COVID-19 inpatient death | 80786 (64.5) | 5391 (41.2) | 88675 (100) | 17 (0) | 17 (0.1) | 0 (0) | 88675 (2.6) |

**Supplement table 5: Primary diagnosis on death certificate for deceased patients**
Table shows the top 10 most frequent primary diagnosis on the death certificate for individuals dying with or without a COVID-19 diagnosis. After removal of duplicates and missing; 125,869 primary diagnosis was found for the 123,648 individuals with COVID-19 on the death certificate, and 12,569 primary diagnosis for the 12,547 dying without COVID-19 on the death certificate within 28 days of a COVID-19 event.

| **COVID on the death certificate** | | | **Without COVID on the death certificate** | | |
|---|---|---|---|---|---|
| **ICD10** | **Description** | **N (%)** | **ICD10** | **Description** | **N (%)** |
| U071 | COVID-19 Confirmed | 109603 (87.5%) | F03 | Unspecified dementia | 890 (6.8%) |
| U072 | COVID-19 Suspected | 3514 (2.8%) | J189 | Pneumonia | 710 (5.4%) |
| F03 | Unspecified dementia | 1218 (1%) | C349 | Cancer of bronchus and lung | 659 (5%) |
| I259 | Chronic ischemic heart disease | 709 (0.6%) | J440 | COPD | 443 (3.4%) |
| C349 | Cancer of bronchus and lung | 685 (0.5%) | I259 | Chronic ischemic heart disease | 436 (3.3%) |
| G309 | Alzheimer's disease | 643 (0.5%) | I64 | Stroke | 421 (3.2%) |
| I64 | Stroke | 639 (0.5%) | G309 | Alzheimer's disease | 388 (3%) |
| I219 | Acute myocardial infarction | 531 (0.4%) | I219 | Acute myocardial infarction | 361 (2.8%) |
| F019 | Dementia | 499 (0.4%) | F019 | Dementia | 309 (2.4%) |
| J440 | COPD | 314 (0.3%) | C61 | Malignant neoplasm of prostate | 283 (2.2%) |
